# Evolution and impact of the strategy to eliminate *gambiense* human African trypanosomiasis in Guinea

**DOI:** 10.64898/2026.03.16.26348467

**Authors:** Moïse Kagbadouno, Ronald E Crump, Samuel A Sutherland, Rob Sunnucks, Oumou Camara, Ching-I Huang, Mamadou B Diallo, Mamady Camara, Favié Béavogui, Dansy Camara, Kevin Allain, Paul E C Brown, Abdoulaye Diaby, Bruno Bucheton, Paul R Bessell, Emily H Crowley, Jean-Mathieu Bart, Kat S Rock, Mamadou Camara

**Affiliations:** Programme National de Lutte contre les Maladies Tropicales Négligées (PNLMTN), Conakry, Guinée; University Gamal Abdel Nasser of Conakry, Guinea; Systems Biology and Infectious Disease Epidemiology Research (SBIDER), University of Warwick, Coventry, UK; Mathematics Institute, University of Warwick, Coventry, UK; Warwick Medical School, University of Warwick, Coventry, UK; Centre for Interdisciplinary Methodologies (CIM), University of Warwick, Coventry, UK; INTERTRYP, Université de Montpellier, CIRAD, IRD, Montpellier, France, Montpellier, France; Epi Interventions Ltd, Edinburgh, UK; Foundation for Innovative New Diagnostics, Geneva, Switzerland

**Keywords:** Sleeping sickness, *gambiense* human African trypanosomiasis, modelling, disease burden, elimination, Guinea, vector control, diagnostics

## Abstract

In 2025 Guinea was validated as achieving elimination as a public health problem for the highly pathogenic, vector-borne infection, *gambiense* human African trypanosomiasis (gHAT) after reaching several years of low-level case reporting. gHAT cases in Guinea have overall seen a large decrease between 2000 and 2024, however there have been notable fluctuations. Transmission modelling was used to assess these trends in observed cases for the foci Boffa East, Boffa West, Dubréka, and Forécariah and evaluate transmission changes. This study quantifies the impact of interruptions due to Ebola and the introduction of new interventions (particularly the rapid diagnostic tests in the passive health system and vector control) in each focus.

The model suggests that transmission of gHAT has fallen 97% (83–100%) between 2000 and 2024, with disease burden measured in disability-adjusted life years (DALYs) reduced by 94% (63–100%). We estimated that Ebola interruptions caused an additional 1,147 gHAT DALYs due to the suspension of gHAT activities, however passive screening improvements and the introduction of vector control likely averted 1,719 and 9,038 DALYs respectively. This study quantifies the impact of Ebola related interruptions on gHAT transmission and disease burden and highlights the success of medical and vector control interventions in Guinea.

## Context

*Gambiense* human African trypanosomiasis (gHAT), also known as sleeping sickness, is a fatal disease caused by a parasite, *Trypanosoma brucei gambiense*, transmitted to humans by the bite of the tsetse fly, which is the vector.

The disease re-emerged in the 1980s and had a large impact on the overall situation in Africa. In 2012, the WHO, building on the efforts of national programmes to combat gHAT, set two main objectives for fighting sleeping sickness: the first was elimination as a public health problem (EPHP) by 2020, measured using a national indicator (<1 case reported per 10,000 people on average for each endemic health district over the last five years) and also continental indicators (a 90% reduction in the risk area compared to 2000–2004, and fewer than 2,000 cases reported across all active foci). The national indicator was achieved by Togo in 2020, followed by nine other countries by the end of 2025 [1]. Since 2017, the number of global cases has fallen below 2,000 per year [2] and since 2022, the WHO has estimated that the global area at risk for gHAT has decreased from 693,700 km² between 2000 and 2004 to 58,221 km² between 2018 and 2022, a difference of 91.6% [3]. The WHO’s second goal is the elimination of transmission by 2030; for a country to verify this elimination, it must be able to prove the absence of new cases for five consecutive years [4].

Guinea is an endemic country for gHAT. Between 2015 and 2024, Guinea reported 549 cases, representing 4.8% of global cases [5]. Thanks to the efforts of the National HAT Control Programme (PNLTHA), established in 2003, and their various partners, the burden of the disease has declined greatly. Although the area at risk in Guinea decreased by only 51% (from 4,300 km² between 2000 and 2004 to 2,108 km² between 2018 and 2022 [3,6]) the number of cases has drastically decreased in recent years, allowing Guinea to celebrate EPHP in January 2025 [7,8].

Historically, sleeping sickness has been prevalent in all regions of Guinea, with foci in both forest and savanna areas. Since its re-emergence, it has become concentrated along the coast in a new, specific, and complex context: the mangrove. Several factors explain the development of the disease in this biotope, notably the rapid demographic and economic growth of Conakry, which has led to the intensive exploitation of the mangrove through rice cultivation, salt extraction, fishing, and the use of mangrove wood.

### Diagnosis

The PNLTHA led the response, which relied on mass medical screenings to diagnose and treat cases. Diagnostic methods at the time consisted of combining a serological test (CATT) with lymph node palpation, followed by microscopic observation of trypanosomes present in blood, lymph node fluid, and/or cerebrospinal fluid (CSF). These tests were progressively improved, particularly with the use of buffy coats during the mini-AECT (mAECT), which increased the test’s sensitivity [9]. From 2013 onward, the introduction of rapid diagnostic tests (RDTs) enabled the implementation of surveillance at the heart of transmission foci, in approximately 100 health structures [10]. This so-called passive screening method was also complemented by active door-to-door screening, also based on the use of RDTs [11] and was generalised after the COVID-19 epidemic from 2020.

### Diagnosis of Stage

The choice of treatment for patients screened as described above depends on the stage of the disease. In the first stage, known as the haemolymphatic stage, the cerebrospinal fluid (CSF) has a white blood cell count (WBC) of ≤5 leukocytes/µl and the no trypanosomes. In the early second stage (meningoencephalitic), the WBC count in the CSF is between 5 and 100 leukocytes/µl, with or without the presence of trypanosomes. Finally, in the advanced second stage, the WBC count in the CSF is greater than 100 leukocytes/µl.

### Treatment

From 2002 to 2011, melarsoprol and pentamidine were the only drugs used. Pentamidine was administered to treat patients in stage 1 and early stage 2, and melarsoprol for patients in advanced stage 2. The introduction of NECT in 2012 represented a major advance in the management of patients with stage 2 disease, so pentamidine was then reserved for stage 1 patients only. A further step was taken with the introduction of fexinidazole (an oral treatment effective in both stages of the disease) in 2021 into the Ministry of Health’s gHAT treatment protocol. Since the introduction of fexinidazole, 30% of patients have benefited from this treatment, while the remaining patients, in advanced stage 2, continue to receive NECT, which is considered more effective in this clinical presentation [12]. Finally, three clinical trials were conducted in Guinea and the Democratic Republic of Congo (DRC) on acoziborole, the first single-dose oral drug effective at all stages of the disease. The results of these trials being very promising and having now been approved by the European Medicines Agency in February 2026, it is expected that it will be introduced in Guinea by 2027 [13,14]

### Vector control

In conjunction with medical control, Guinea’s PNLTHA focused early on entomological control aimed at cutting the principal transmission link through the reduction of human-vector contact. Between 2006 and 2008, vector control (VC) methods and techniques against tsetse were evaluated on the Loos Islands [15,16] to select the most appropriate control strategy. The use of impregnated targets proved to be the most effective and environmentally friendly. This technique was subsequently improved by the use of smaller targets, which are less expensive and more effective [17].

In 2012, to assess its impact on transmission in endemic foci, VC via the deployment of Tiny Targets was introduced in the eastern part of the active focus in Boffa, and not in the western part, in combination with screening and treatment of gHAT cases. The results of this study showed that the contribution of VC in combination with medical control had a significant impact on the disease and its incidence [18].

### Ebola

Between 2013 and 2016, the Ebola epidemic struck Guinea. During this period, medical activities for gHAT had to be halted, and only so-called “passive” screening, implemented in health centres thanks to the arrival of RDTs, allowed for the continued detection of a few cases. When fieldwork resumed in 2016–2017, an uptick in the number of cases was observed in all three foci, with the exception of Boffa East, the area where VC had been implemented. In this area, protected from tsetse, no cases were diagnosed. This confirmed that, even in the absence of medical intervention, VC was capable of interrupting transmission [19]. Thanks to these results, the WHO has since suggested integration of VC can support the fight against gHAT. Starting in 2016, this strategy was therefore extended to other active foci in the country (Boffa West and Dubréka in 2016 and Forécariah in 2018). Since 2018, more than 15,000 Tiny Targets have been deployed annually, with increasing community participation. These deployments have led to a large decrease in tsetse density and consequently a reduction in the risk of transmission [20]. This strategy has strongly contributed to achieving the objective of EPHP of gHAT in Guinea in January 2025 [7,8].

In this article, we describe the specific changes made to strategies to control gHAT in Guinea between 2000 and 2024. We use mathematical modelling to estimate the impact of these strategies on transmission and case reporting in the foci that remain endemic in the country (Boffa, Dubréka and Forécariah). We use medical and vector data that are fed into our mathematical models to try to understand the evolution of the disease in Guinea over the last 25 years.

Following the fitting of our model to the data, we used it to perform a counterfactual analysis. Our model was used to quantify the impact of the 2013–2016 Ebola epidemic, the introduction of RDTs into passive screening, and the contribution of Tiny Target deployments. The actual scenario was compared to three counterfactual scenarios: no Ebola outbreak, no improvement in passive screening, and no Tiny Target deployments. Through these comparisons, we were able to estimate their effects on case detection, infection dynamics, and the burden of disease measured in disability-adjusted life years (DALYs).

We present our results in this article for each focus separately, as well as at the national level. We discuss the main transmission factors based on the estimation of model parameters such as the relative risk of tsetse exposure, evidence of animal transmission, passive detection rates, and the contribution to transmission of asymptomatic human skin infections.

This is the first time the impact of VC, improved passive screening, and the Ebola epidemic on gHAT have been quantified for Guinea using a model. It is important to estimate what happened to transmission and DALYs, which are not directly observable. These results are also available in our interactive graphical interface (https://hatmepp.warwick.ac.uk/guineafitting/v4/).

## Methods

### Data used for the modelling

#### Demographic data

The at-risk population in each focus area was estimated for 2024 at 20,463 in Boffa West, 5,639 in Boffa East, 30,693 in Dubréka, and 22,132 in Forécariah. The values for Boffa West and Boffa East are from intensive population surveys conducted in 2011 [18], the value for Forécariah is from a population survey conducted in 2008 [21], and the value for Dubréka is based on an estimate made by the PNLTHA in 2016. These values were adjusted year by year using the annual population growth rate of the administrative unit containing the focus of the disease (see Table A of the SI), assuming exponential growth.

#### Medical data

Data for the period 2000–2022 were obtained from the WHO HAT Atlas [3,22] and aggregated into annual values within each focus using geolocation information where available and place names where not available. The PNLTHA provided aggregated data for the period 2023–2024 (see the SI Excel file). The data used included the number of people actively tested, the number of cases detected through active screening, and the number of cases detected through passive screening. Detected gHAT cases were classified by disease stage where this information was available.

The number of people tested during the Ebola period under passive screening [23] was used to determine when passive screening was improved and interrupted by Ebola (Figure A of the SI).

#### Entomological data

We use the number of Tiny Targets deployed during each deployment to inform our projections and adjustments to the tsetse model. Tsetse density monitoring is carried out through entomological assessments at eco-epidemiologically defined sentinel traps to evaluate the impact of the VC on apparent trap densities (ATDs) (the number of flies per trap per day). This monitoring takes place three to four months after the deployment of the Tiny Targets in the field. Across the four intervention zones of the three active foci, 91 biconical sentinel traps were used (20 in Boffa West, 20 in Boffa East, 20 in Dubréka, and 31 in Forécariah) [24]. This entomological monitoring was initially carried out at a frequency of three entomological assessments per year (around March, May and November) in the pilot phase of the VC (2012–2015), two assessments per year (around May and October) in the generalisation phase (2016–2022), and once a year (around May) since 2023 (see the Excel file in the SI for more information on these deployments).

### Tsetse and HAT models and their adjustment

#### Tsetse model

The compartmental ordinary differential equation (ODE) model for tsetse describes pupae that emerge as adults susceptible to gHAT infection. If their first blood meal comes from an infected host, they can become infected. In the model, we assume that after the extrinsic incubation period, infected tsetse become infectious and remain so for life. If they do not become infected, they will have reduced susceptibility to gHAT during subsequent blood meals (the teneral effect [25]).

To simulate the impact of Tiny Targets, we include a parameter that increases the mortality rate after deployments, but this parameter decreases after a few months due to several factors, such as Tiny Target detachment, disappearance, or immersion due to tides [24]. See the SI text for more detailed information on this model.

The tsetse model was fitted to the ATD at each monitoring time, taking into account the number of Tiny Targets used during each deployment, using maximum likelihood estimation. The estimated parameters were (i) the probability that a fly would contact a Tiny Target and die (per target), (ii) the tsetse birth rate (which controls the recovery of the fly population between deployments), (iii) the intercept of the population size before VC, and (iv) the level of tsetse re-emergence (only in Boffa East).

The estimates of the tsetse model parameters were included as fixed values when fitting the gHAT transmission model. This allowed the model to account for differences in VC performed in each focus, and in particular for variations in the number of Tiny Targets deployed over time.

#### Transmission model

We used historical case reporting data and the number of people tested during active screening from the WHO HAT Atlas (2000–2022), as well as information and data related to the implementation of gHAT control activities by the Guinea National HAT Control Programme (PNLTHA-Guinea) (2023–2024). Using this information, we fitted our previously developed transmission model to these data [27]. We do not fit our model to data from historical foci within the country. We used nine variants of the model, some of which account for the heterogeneity of risk from tsetse bites, others for animal transmission, and another variant for asymptomatic human skin infections. Using statistical weighting, we use the adjustments from these models to produce an ensemble model that accounts for the uncertainty of cryptic transmission routes.

The gHAT transmission model is a compartmental model. Several variants exist, defined in terms of the human population structure (high or low risk of infection, and participating in active screening randomly or not at all) [27,28]. It also considers the possible contribution to transmission from asymptomatic animal or human infections [26,29] (see the equations in Section S1.3 and Table B of the SI for the definition of the nine model variants considered). Both deterministic and stochastic versions of the transmission model exist. The deterministic version is used in fitting the model to the data for computational speed, while the stochastic version is used in model simulations to better represent stochastic effects [30].

The deterministic gHAT transmission model was fitted to the historical case data [27] with an adaptive Markov chain Monte Carlo (MCMC) statistical approach [27,29].

This model accounts for improvements in the passive screening system and the variable number of people screened each year, including during the Ebola epidemic. Parameters expected to vary geographically were estimated during this fitting process. These included (i) the basic reproduction number of the disease, (ii) the proportion of the population according to their risk level for infection and participation in active screening, (iii) the relative sizes of passive screening improvements, and (iv) parameters associated with transmission through asymptomatic animal or human infections (see Table E of the SI for a complete list).

For the model including asymptomatic human transmission and self-cure, we assume that the parameters are intrinsic biological values and are not subject to local geographic variations. These parameters were (i) the proportion of human exposures resulting in an initial bloodstream infection, (ii) the self-cure rates for stage 1 bloodstream infections, (iii) the self-cure rate for skin infections, (iv) the rate of transition from a skin infection to a bloodstream infection, and (v) the relative infectivity of skin infections compared to bloodstream infections. Because the parameters in this model are not expected to vary geographically, but we fit them to the data from each focus separately, we used sequential Bayesian updating to share information from the most informative focus to the least informative one [26].

### Counterfactual scenarios

We simulated our model with several counterfactual scenarios (CFS) that show the different potential trajectories of gHAT if the past situation had been different. These three CFS are described in Table 1. For these simulations, we adapted a version of the simulation method that allows for more robust comparisons between the different scenarios (See the SI for more information) [31].

**Table 1.**
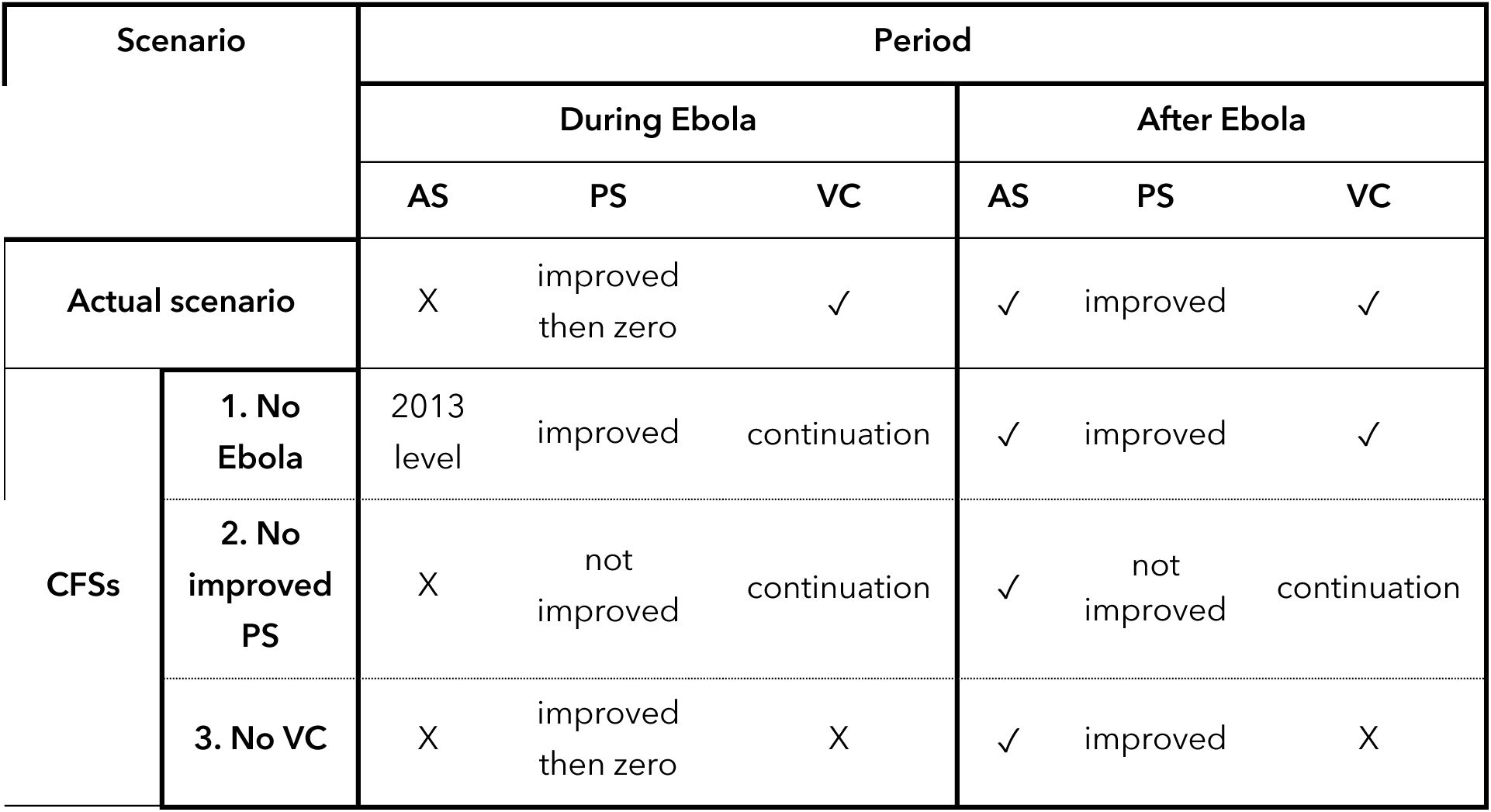
Actual and counterfactual scenarios considered. Each row shows a scenario: the first is the actual scenario that took place and includes no active screening (AS) during the Ebola period, initial improvements in passive screening (PS) at the start of the Ebola period then interruption of PS. After Ebola, AS and PS resumed. In the actual scenario, vector control (VC) continued during Ebola where it had already started, and was implemented in all foci after Ebola. The other three rows show the components added or removed to the actual scenario to make up our counterfactual scenarios (CFSs): 1. No Ebola, 2. No PS improvements, or 3. No VC.

| Scenario |  | Period |  |  |  |  |  |
| --- | --- | --- | --- | --- | --- | --- | --- |
|  |  | During Ebola |  |  | After Ebola |  |  |
|  |  | AS | PS | VC | AS | PS | VC |
| Actual scenario |  | X | improved then zero | ✓ | ✓ | improved | ✓ |
| CFSs | 1. No Ebola | 2013 level | improved | continuation | ✓ | improved | ✓ |
|  | 2. No improved PS | X | not improved | continuation | ✓ | not improved | continuation |
|  | 3. No VC | X | improved then zero | X | ✓ | improved | X |

## Results

### Model fitting

#### Tsetse model

Figure 3 illustrates our tsetse model fit to the ATD data. It shows how, in a context of substantial decline in tsetse density after target deployment, this relatively simple model can capture the overall dynamics observed during entomological surveillance. It was important to simulate the variable number of Tiny Targets deployed in each focus each year, as the fits were less good at capturing the trends if we assumed constant deployment each year. Similarly, taking into account the re-emergence of tsetse in Boffa East in 2018 improved our fit to the trap data.

**Figure 1.**
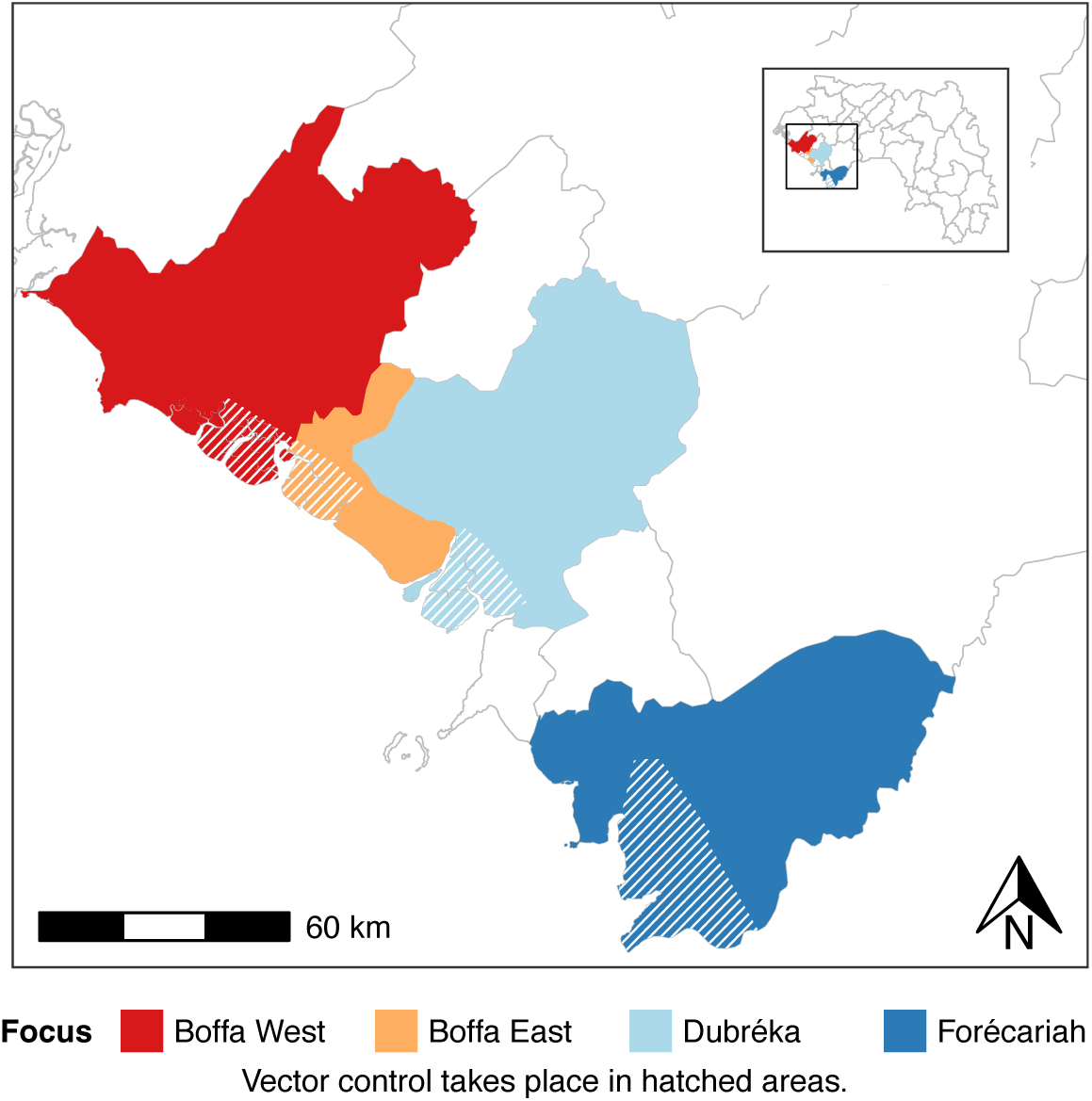
Map of foci. The boundaries correspond to the prefectures or sub-prefectures (in the case of Boffa East) where the foci are located. Vector control is carried out in the hatched areas contained within each focus.

**Figure 2.**
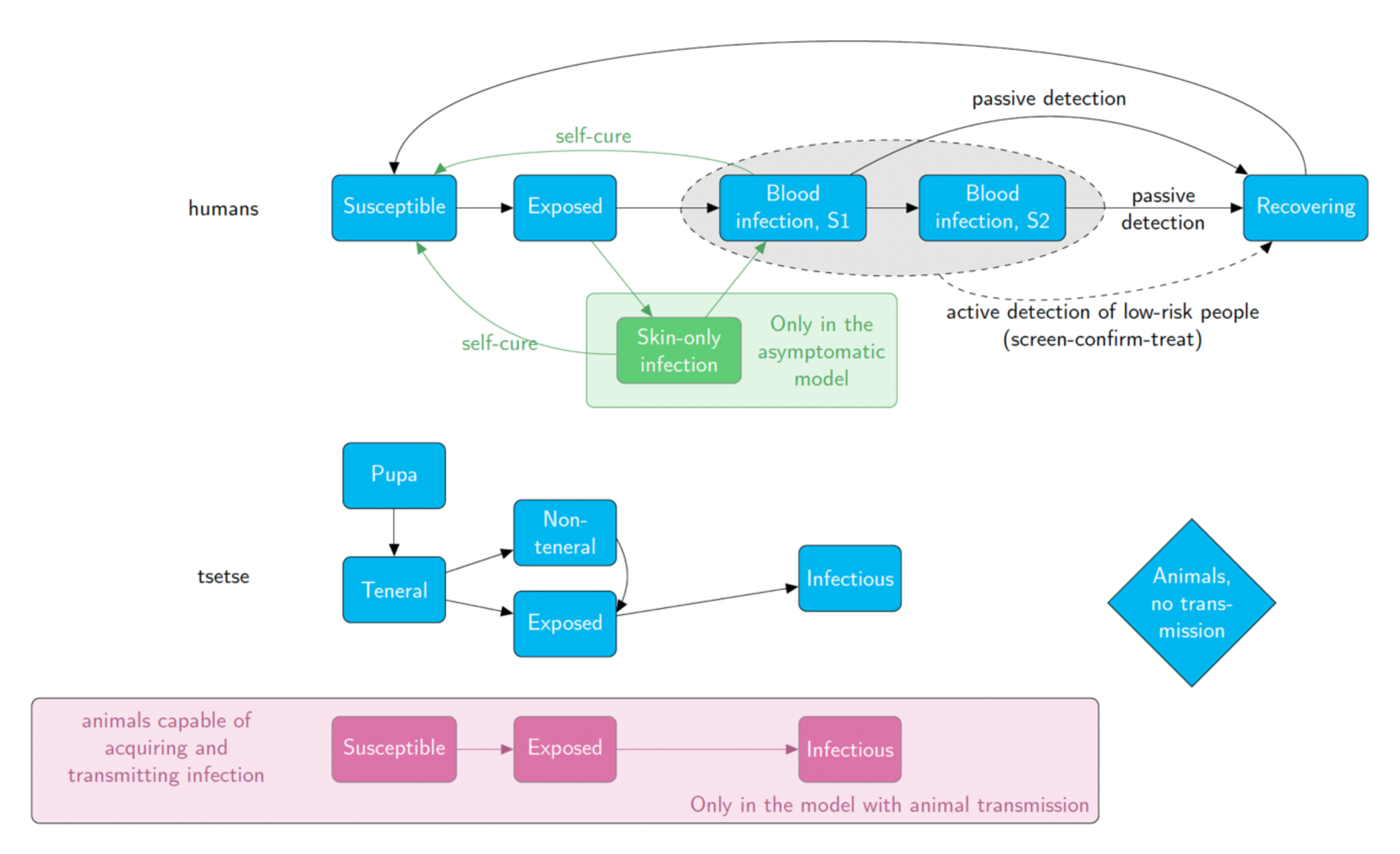
Schematic of our compartmental model and its variants. The top line corresponds to the progression of gHAT infection in humans from susceptible through different stages of infection and then to recovered. Similar boxes are shown for infection in tsetse. The pink boxes show compartments that are included in our model with animal infections, and green boxes and arrows show the one extra compartment for humans with skin-only infections included in the variant with asymptomatic transmission. Corresponding equations and parameterisation for the model is presented in the SI (reproduced under a CC-BY license from Crump et al. [26]).

**Figure 3.**
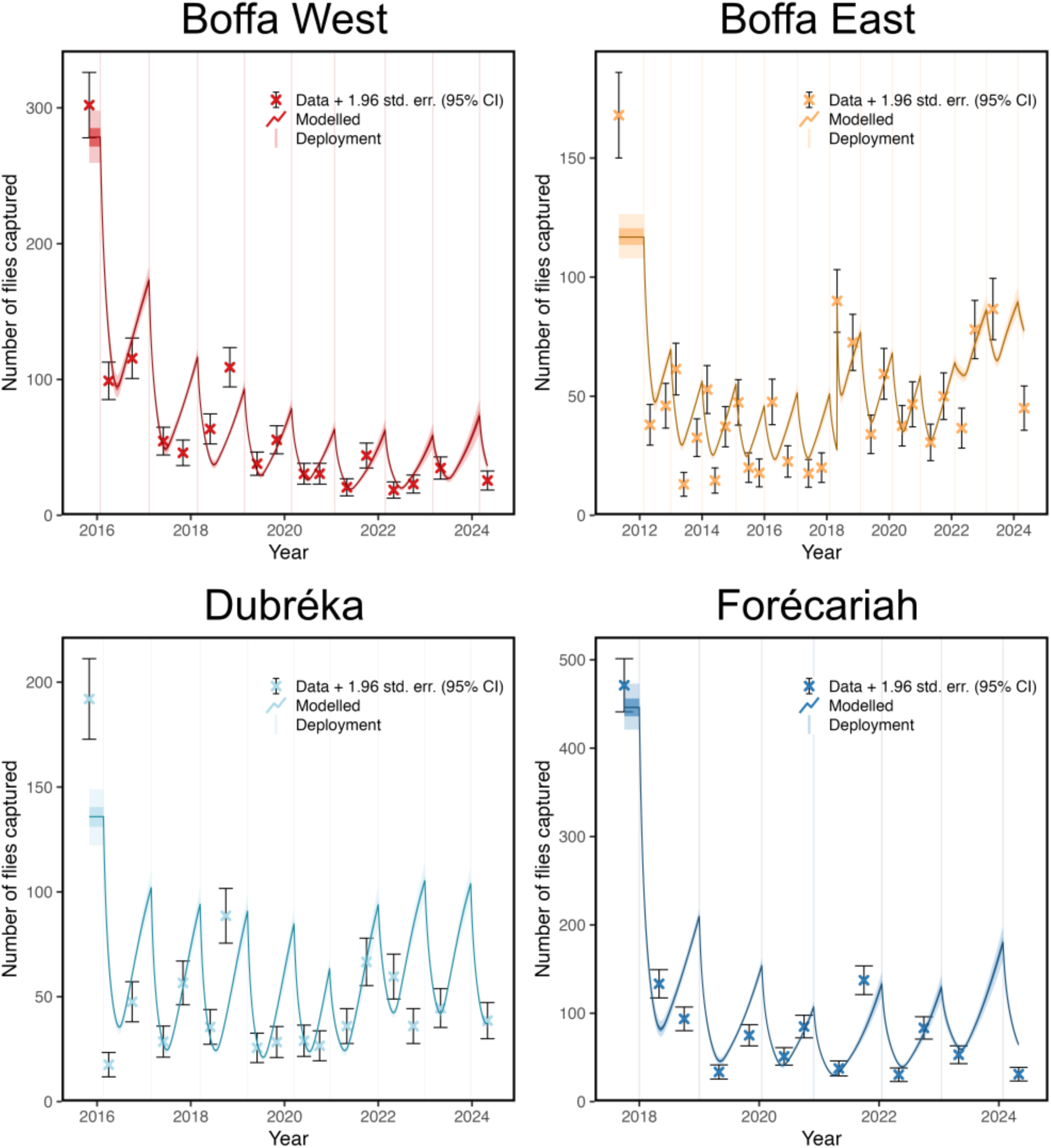
Tsetse model fit to trap data from each focus. Data points with error bars indicate the observed value and Poisson error. The line indicates the best fit, and the shaded areas indicate the 50% and 95% confidence intervals for the model (using the maximum likelihood estimation (MLE) statistical method for the fitting and the inverse of the hessian matrix for the confidence intervals).

In the Dubréka and Boffa East foci, there is a clear difference between the model and the data for the first monitoring point (pre-deployment of Tiny Targets) as well as for a few other points where variability is not as well represented. However, in Boffa West and Forécariah, there is a good match between the tsetse model and the data. For all foci, the model successfully took into account the decline in densities over time, as well as the gradual malfunctioning of the tiny targets during the year, illustrated by the periodic peaks in the model.

#### Epidemiological model

Our ensemble model is comprised of a weighted combination of the nine model variants. Statistically comparing each variant’s fit to the data in each location, we found that Models 4, 7 and 9 were well supported by the data; all of these model variants had a structure with heterogeneity in risk of exposure to tsetse bites and high-risk people not participating in AS, with Model 7 having additional animal transmission and Model 9 having additional skin-only infections in humans (see Table G in the SI). This suggests that it is very important to capture the high/low-risk structure in our model to match the observed trends in case reporting, and whilst animals or asymptomatic transmission could be driving transmission, it is uncertain. Boffa East had the most support for the hypothesis that asymptomatic transmission is occurring and Dubréka had the least.

Figure 4 presents the ensemble model’s predictions regarding the number of cases recorded during active and passive screening, as well as the annual number of new infections for the study period, compared to the observed data for each of the four foci. To represent the parameter uncertainty generated by the model, we generated a total of 20,000 simulations for each focus which are represented by the box and whisker plots. In all four foci, the agreement between the model and the observed data is good, as shown by the black line, and the ensemble model results are represented by the coloured box and whisker plots (Figure 4). The impact of VC on gHAT transmission is reflected in a rapid decrease in the number of new infections in the years this control programme was implemented in each focus.

**Figure 4.**
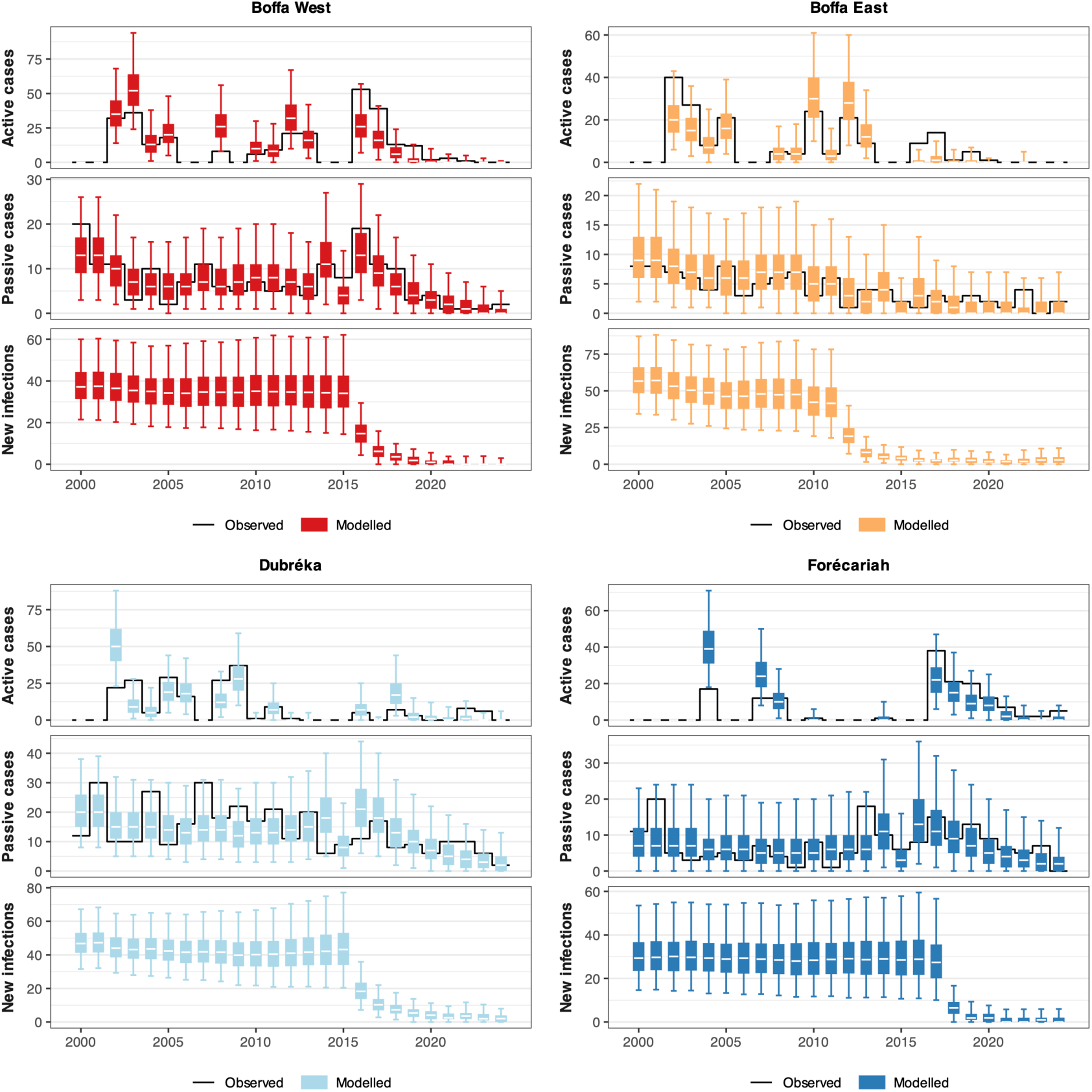
Adaptation of the transmission model to the epidemiological data for each focus. We show a panel of four fits, one for each focus. The observed data can be seen as black lines, and the ensemble model results as box plots. For each fit, the first row represents active cases, the second passive cases, and the third new infections estimated by the model (but these are not directly observable).

Between 2000 and 2024 we estimate that the number of new infections fell 97% (83–100%) from around 171 (134–236) per year across all foci in 2000 to 3 (0–11) per year in 2024. Similarly, the model suggests that disease burden has dropped from 3,785 (2,587–5,125) DALYs per year in 2000 to 3 (0–235) DALYs per year in 2024.

### Counterfactual Scenarios

Figure 5 and Table 2 compare the actual scenario with the three CFSs: 1. No Ebola, 2. No improvement to PS and 3. No VC, over the period 2012–2024 (cumulative results). We selected this period as 2012 is the first year where there was VC in Guinea (in Boffa East) and we wanted to be able to make fair comparisons (with the same year ranges) between the different scenarios and foci. If no Ebola outbreak had occurred (CFS 1 in Table 2), the model finds that more cases would have been detected, there would have been fewer new infections and deaths due to gHAT, and fewer DALYs in all foci throughout the period. Without improvement to PS from early 2014 (CFS 2 in Table 2), the model suggests there would have been fewer cases being detected and more new infections, deaths, and DALYs. Without VC (CFS 3 in Table 2), new infections, deaths and DALYs would have all been higher. The model estimated that VC averted 9,038 DALYs (95% prediction interval (PI): 3,241–20,365) across all foci when compared to the actual scenario. The implementation of VC was calculated to have averted more DALYs compared to improved PS (1,719 DALYs, 95% PI: 300–4,360) or compared to those gained due to the Ebola outbreak (1,147 DALYs, 95% PI: 294–2,515), see Table H in the SI for the 95% prediction intervals of the 1,012–2,024 cumulative results.

**Figure 5:**
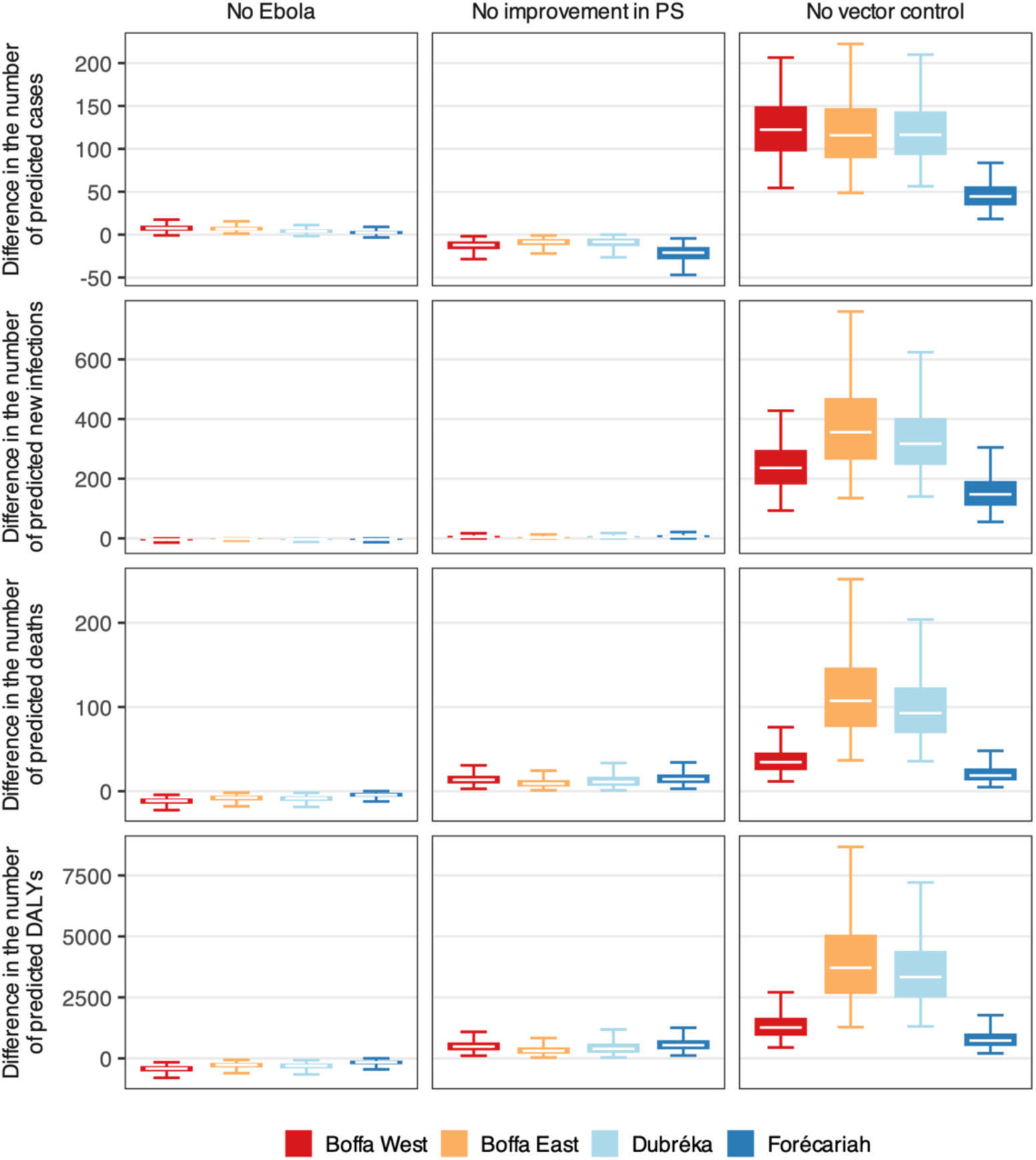
Impact of the Ebola epidemic, improvements in passive screening and vector control (VC) over the period 2012–2024. These impacts are represented by the difference between the number of reported cases, new infections, deaths and disability-adjusted life years (DALYs) due to gHAT in the actual scenario and in each counterfactual scenario (1–3).

**Table 2.**
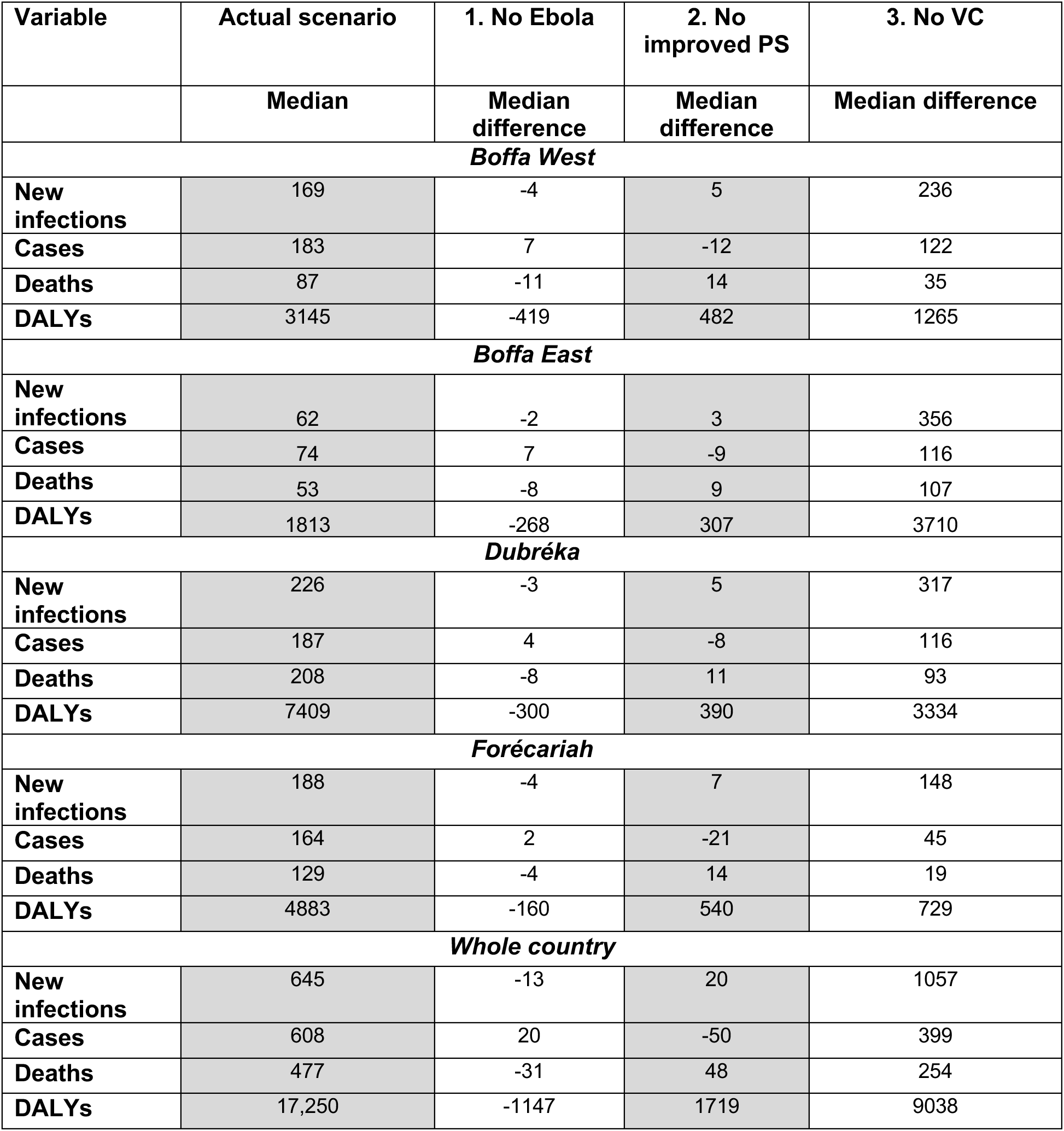
Model estimates for the number of new infections, cases, deaths due to gHAT and DALYs under the actual scenario during the period 2012–2024 (second column) and the impact of the Ebola epidemic, improved passive screening (PS), and vector control (VC) (third to fifth columns) in terms of difference in expected new infections, cases, deaths and DALYs.

Time series trends in case reporting and new infections under the actual scenario and the three CFSs for all foci can be found in Figures D to G in the SI.

## Discussion

There has been limited prior use of modelling to assess past gHAT strategies in Guinea. Ten years ago, Pandey et al. [32] used compartmental transmission model variants (with and without animal transmission) and fitted them to four years of active screening data from Boffa. Although, they did not assess which of the two models was best supported by the data, they did find that more sustained vertical interventions would be needed if animal transmission was occurring. Their study focused on future strategies to achieve EPHP in the focus and it therefore did not directly evaluate the effectiveness of past intervention. A subsequent paper used a different approach (parameterised by one year point prevalence data, but including unconfirmed seropositives), which concluded that to maintain gHAT infection in Forécariah, asymptomatic infections that contribute to transmission would be needed if no transmission was occurring through animals [33]. In the present study, the conclusion was similar: our model variants required some type of host heterogeneity to match the case reporting trends observed in all foci of Guinea and also found support for Model 9 which had asymptomatic human transmission. This finding contrasts with modelling analysis of asymptomatic transmission for the DRC which found very low statistical evidence for asymptomatic transmission [26] and raises a question about whether the West African human-trypanosome interaction is different to that in Central Africa.

Camara et al. previously evaluated the likely impact on gHAT disease burden due to Ebola in Guinea [23]. That analysis used a different approach to the present study, taking the reported cases and deaths before and during the Ebola period to extrapolate DALYs for the country. They concluded that DALYs during Ebola were around 3.5–9.6 times higher than before the outbreak due to reduced medical capacity to diagnose and treat gHAT. Finally, Kagbadouno et al. assessed the role of VC during Ebola on case reporting finding that whilst case reporting after Ebola increased to a prevalence higher than before the outbreak in 2016 for foci without VC, in Boffa East, which had had VC, there were no cases reported in 2016 [19].

The present study reinforces the message presented in these two studies–interruption of medical activities (in particular passive screening) led to an increase in DALYs compared to what would have been expected if the Ebola outbreak had not occurred, however, VC was able to reduce cases and DALY burden in Boffa East despite Ebola. Other modelling has explored interruptions to gHAT programmes due to other causes, echoing the findings presented here. For example, Huang et al. found that, in the DRC, interruption of active screening for a short period had a negative but relatively small impact [34]. However, maintaining functional passive screening during any interruption was very important for short to mid-term impact on disease burden and, for locations that had already implemented VC, there was a protective effect.

CFSs are a good way to calculate the impact on transmission and disease burden and were also used in Mandoul (Chad) where there was no control area to compare to the intervention area [35]. As Courtin et al. [18] already demonstrated the value of the entomological approach in Boffa East and West it made practical and ethical sense to implement VC in all foci in Guinea.

### Limitations

In this article we have focused on the assessment of past strategies that were put in place in Guinea and used the transmission model to quantify progress towards elimination and impact of the different intervention components. We have not simulated potential future strategies in the present study. However, the calibrated model presented here provides a robust baseline from which different strategy options could be evaluated for their ability to reach elimination of transmission and their cost-effectiveness. Subsequent work by the authors will use this approach to support future decision making.

During 2022–23 Guinea took part in a safety and tolerability study to assess the new drug, acoziborole, in the treatment of unconfirmed seropositives [36]. The treatment of the seropositives enrolled in this study was not included in our model, however, if any of these treated people were contributing to transmission to tsetse or progressed to become confirmed cases, there would have been a reduction in transmission and/or case reporting as a result. As the use of this “screen-and-treat” approach was only used for a short period and not for all seropositive individuals, it is unlikely that this omission will have led to a large difference in the model outcomes.

Our model considered each focus as a separate, fixed population with the same populations of humans and flies mixing throughout the year. We assumed no movement between the foci, nor movement of people into and out of the foci. This simplification was made to avoid introducing additional complexity into the model; this would have resulted in a more computationally intensive fitting procedure as well as the introduction of several unknown parameters, many of which would likely be statistically unidentifiable. In practice, we note that there is movement of populations, particularly seasonal movements associated with certain livelihoods (e.g. salt production, rice growing, and fishing).

In recent years there has been an apparent emergence of a new peri-urban focus of cases in Dubréka. In this analysis we considered all cases in Dubréka to be part of the same focus, however it is possible that transmission in this peri-urban location is an epidemiologically distinct focus. Separating out the historically endemic and peri-urban foci in Dubréka, would have resulted in too few data points to fit our model to both areas, however, future modelling analyses should consider if this is possible and whether the new focus requires a new or different strategy compared to the historical focus.

## Conclusions

This study provides a quantitative measurement of the success in the fight against gHAT in Guinea in the last decades. Our transmission model for gHAT in Guinea has estimated that there has been a 96% reduction in new infections and a 92% reduction in DALY burden between 2000 and 2024 thanks to a combination of medical interventions (including new diagnostic tools) and VC.

VC, in particular, is found by the model to have had a considerable impact on transmission. All endemic foci in Guinea have had VC in place since at least 2018 resulting in substantial reductions in the tsetse populations. The model results indicate that this activity has greatly reduced new infection events. Our counterfactual scenario, modelling what we would have expected without VC, finds 1,057 (95% PI: 423–2,118) more new infections in Guinea between 2012–2024 compared to the 645 (95% PI: 326–1,145) new infections estimated to have actually occurred. This equates to around 62% fewer infections due to VC.

Improvements to passive screening, through the introduction of rapid diagnostic tests were estimated to have reduced DALYs. Without passive screening improvements, the counterfactual model computed there would have been around 1,719 (95% PI: 300–4,360) more DALYs compared to the 17,250 (95% PI: 8,542–30,771) DALYs between 2012–2024 in the actual scenario. Conversely, the Ebola outbreak increased DALY burden in Guinea due to gHAT intervention interruptions with an increase of 1,147 (95% PI: 294–2,515) DALYs; this would have been far worse if improved PS had not been implemented after the outbreak and if VC had not continued in Boffa East.

Guinea has overcome a high level of gHAT prevalence and obstacles such as the West African Ebola outbreak. However, the work does not stop here and continued efforts will be needed to capitalise on the success of the programme and terminate gHAT in Guinea for good.

## Supporting information

Epidemiological data

Vector control data

## Data Availability

All data used are available as supplementary data files and as part of an OSF, which also contains the results produced.

https://doi.org/10.17605/OSF.IO/97PS6

## Acknowledgements

The authors would like to thank Prof Simon Spencer for his advice concerning the statistical methodology for model fitting. We would also like to thank the WHO for collation and access to epidemiological data in the framework of the HAT Atlas [3].

## Author contributions

- Conceptualisation KSR, MC, REC
- Methodology REC, RS, SAS
- Software PEB
- Validation
- Formal Analysis REC, CH, SAS
- Investigation
- Data Curation MK, OC
- Writing - Original Draft MK, REC, OC, JMB, MC, KSR, SAS, KA
- Writing Review and editing all authors
- Visualisation REC, PEB, KA
- Supervision KSR, JMB, MC
- Project administration EHC, MC
- Funding acquisition KSR, EHC,

## Data and code

Aggregated information about the WHO HAT Atlas data used for fitting is described in the Supplementary Information. Original data cannot be shared publicly because they are under the stewardship of the World Health Organization’s HAT Atlas; our data-sharing agreement does not allow us to share that data. Original data are available from the WHO (contact or visit https://www.who.int/health-topics/human-african-trypanosomiasis/) for researchers who meet the criteria for access to confidential data. PNLTHA data used for this model fitting analysis is also available in the Supplementary Information. All modelled outputs and code are available via Open Science Framework: https://doi.org/10.17605/OSF.IO/97PS6

## Supplementary material

S1 Text – Additional methodology and results

S2 Text – French version of main text

S1 File – VC data

S2 File – Epidemiological data

## References

1. World Health Organisation. 2025 https://www.who.int/news/item/08-08-2025-kenya-achieves-elimination-of-human-african-trypanosomiasis-or-sleeping-sickness.

2. Franco JR, Cecchi G, Paone M, Diarra A, Grout L, Kadima Ebeja A, Simarro PP, Zhao W, Argaw D. 2022 The elimination of human African trypanosomiasis: Achievements in relation to WHO road map targets for 2020. PLoS Negl. Trop. Dis. 16, e0010047-.

3. Franco JR, Priotto G, Paone M, Cecchi G, Ebeja AK, Simarro PP, Sankara D, Metwally SBA, Argaw DD. 2024 The elimination of human African trypanosomiasis: Monitoring progress towards the 2021–2030 WHO road map targets. PLoS Negl. Trop. Dis. 18, e0012111-.

4. World Health Organization. 2023 Criteria and procedures for the verification of elimination of transmission of T. b. gambiense to the human population in a given country.

5. World Health Organisation. In press. The Global Health Observatory. https://www.who.int/data/gho/data/indicators/indicator-details/GHO/hat-tb-gambiense. See https://www.who.int/data/gho/data/indicators/indicator-details/GHO/hat-tb-gambiense (accessed on 12 December 2025).

6. Simarro PP, Cecchi G, Franco JR, Paone M, Diarra A, Ruiz-Postigo JA, Fèvre EM, Mattioli RC, Jannin JG. 2012 Estimating and Mapping the Population at Risk of Sleeping Sickness. PLoS Negl. Trop. Dis. 6, e1859-.

7. World Health Organization. 2025 https://www.who.int/news/item/29-01-2025-guinea-eliminates-human-african-trypanosomiasis-as-a-public-health-problem. https://www.who.int/news/item/29-01-2025-guinea-eliminates-human-african-trypanosomiasis-as-a-public-health-problem.

8. Kagbadouno M, et al. 2026 Elimination of gambiense human African trypanosomiasis as a public health problem in Republic of Guinea: paving the way for zero transmission. Under review

9. Camara M, Camara O, Ilboudo H, Sakande H, Kaboré J, N’Dri L, Jamonneau V, Bucheton B. 2010 Sleeping sickness diagnosis: use of buffy coats improves the sensitivity of the mini anion exchange centrifugation test. Tropical Medicine & International Health 15, 796–799. (10.1111/j.1365-3156.2010.02546.x)

10. Camara O, Biéler S, Bucheton B, Kagbadouno M, Mathu Ndung’u J, Solano P, Camara M. 2021 Accelerating elimination of sleeping sickness from the Guinean littoral through enhanced screening in the post-Ebola context: A retrospective analysis. PLoS Negl. Trop. Dis. 15, e0009163. (doi:10.1371/journal.pntd.0009163)

11. Camara O et al. 2024 Conducting active screening for human African trypanosomiasis with rapid diagnostic tests: The Guinean experience (2016–2021). PLoS Negl. Trop. Dis. 18, e0011985. (doi:10.1371/journal.pntd.0011985)

12. World Health Organization. 2024 Guidelines for the treatment of human African trypanosomiasis. https://www.who.int/publications/i/item/9789240096035.

13. European Medicines Agency. 2026 https://www.ema.europa.eu/en/news/new-single-dose-oral-treatment-human-african-trypanosomiasis-sleeping-sickness. *European Medicines Agency*.

14. ClinicalTrials.gov. 2025 Efficacy and Safety of Acoziborole (SCYX-7158) in Patients With Human African Trypanosomiasis Due to T.b. Gambiense (OXA002). https://clinicaltrials.gov/study/NCT03087955?tab=results.

15. Kagbadouno M, Camara M, Bouyer J, Hervouet JP, Courtin F, Jamonneau V, Morifaso O, Kaba D, Solano P. 2009 Tsetse elimination: its interest and feasibility in the historical sleeping sickness focus of Loos islands, Guinea. Parasite 16, 29–35.

16. Kagbadouno MS, Camara M, Bouyer J, Courtin F, Onikoyamou MF, Schofield CJ, Solano P. 2011 Progress towards the eradication of Tsetse from the Loos islands, Guinea. Parasit. Vectors 4, 18. (doi:10.1186/1756-3305-4-18)

17. Rayaisse JB et al. 2011 Towards an Optimal Design of Target for Tsetse Control: Comparisons of Novel Targets for the Control of Palpalis Group Tsetse in West Africa. PLoS Negl. Trop. Dis. 5, e1332. (doi:10.1371/journal.pntd.0001332)

18. Courtin F et al. 2015 Reducing Human-Tsetse Contact Significantly Enhances the Efficacy of Sleeping Sickness Active Screening Campaigns: A Promising Result in the Context of Elimination. PLoS Negl. Trop. Dis. 9, e0003727. (doi:10.1371/journal.pntd.0003727)

19. Kagabadouno M et al. 2018 Ebola outbreak brings to light an unforeseen impact of tsetse control on sleeping sickness transmission in Guinea. bioRxiv , 202762. (doi:10.1101/202762)

20. Camara AD et al. 2026 Xeno-monitoring the impact of vector control on trypanosome transmission in the Forecariah sleeping sickness focus (Guinea). Under review

21. Courtin F, Jamonneau V, Camara M, Camara O, Coulibaly B, Diarra A, Solano P, Bucheton B. 2010 A geographical approach to identify sleeping sickness risk factors in a mangrove ecosystem. Tropical Medicine & International Health 15, 881–889. (10.1111/j.1365-3156.2010.02559.x)

22. Simarro PP et al. 2010 The Atlas of human African trypanosomiasis: a contribution to global mapping of neglected tropical diseases. Int. J. Health Geogr. 9, 57. (doi:10.1186/1476-072X-9-57)

23. Camara M et al. 2017 Impact of the Ebola outbreak on Trypanosoma brucei gambiense infection medical activities in coastal Guinea, 2014-2015: A retrospective analysis from the Guinean national Human African Trypanosomiasis control program. PLoS Negl. Trop. Dis. 11, e0006060. (doi:10.1371/journal.pntd.0006060)

24. Kagbadouno MS et al. 2024 Population genetics of Glossina palpalis gambiensis in the sleeping sickness focus of Boffa (Guinea) before and after eight years of vector control: no effect of control despite a significant decrease of human exposure to the disease. Peer Community Journal 4. (doi:10.24072/pcjournal.383)

25. Van Hoof L, Henrard C, Peel E. 1937 Influences modificatrices de la transmissibilité cyclique du Trypanosoma gambiense par Glossina palpalis. Ann Soc Belg Med Trop 17, 249–272.

26. Crump RE et al. 2024 Modelling timelines to elimination of sleeping sickness in the Democratic Republic of Congo, accounting for possible cryptic human and animal transmission. Parasit. Vectors 17, 332. (doi:10.1186/s13071-024-06404-4)

27. Antillon M et al. 2024 Cost-effectiveness of end-game strategies against sleeping sickness across the Democratic Republic of Congo. medRxiv , 2024.03.29.24305066. (doi:10.1101/2024.03.29.24305066)

28. Rock KS, Torr SJ, Lumbala C, Keeling MJ. 2015 Quantitative evaluation of the strategy to eliminate human African trypanosomiasis in the Democratic Republic of Congo. Parasit. Vectors 8, 532. (doi:10.1186/s13071-015-1131-8)

29. Crump RE, Huang C-I, Knock ES, Spencer SEF, Brown PE, Mwamba Miaka E, Shampa C, Keeling MJ, Rock KS. 2021 Quantifying epidemiological drivers of gambiense human African Trypanosomiasis across the Democratic Republic of Congo. PLoS Comput. Biol. 17, e1008532. (doi:10.1371/journal.pcbi.1008532)

30. Davis CN, Crump RE, Sutherland SA, Spencer SEF, Corbella A, Chansy S, Lebuki J, Miaka EM, Rock KS. 2024 Comparison of stochastic and deterministic models for gambiense sleeping sickness at different spatial scales: A health area analysis in the DRC. PLoS Comput. Biol. 20, e1011993. (doi:10.1371/journal.pcbi.1011993)

31. Sunnucks R, Davis EL, Rock KS. 2025 Methods for Reproducible Comparison of Strategies in Stochastic Modelling. medRxiv, 2025.10.09.25337145. (doi:10.1101/2025.10.09.25337145)

32. Pandey A, Atkins KE, Bucheton B, Camara M, Aksoy S, Galvani AP, Ndeffo-Mbah ML. 2015 Evaluating long-term effectiveness of sleeping sickness control measures in Guinea. Parasit. Vectors 8, 550. (doi:10.1186/s13071-015-1121-x)

33. Capewell P et al. 2019 Resolving the apparent transmission paradox of African sleeping sickness. PLoS Biol. 17, e3000105. (doi:10.1371/journal.pbio.3000105)

34. Huang C-I, Crump RE, Crowley EH, Hope A, Bessell PR, Shampa C, Mwamba Miaka E, Rock KS. 2023 A modelling assessment of short- and medium-term risks of programme interruptions for gambiense human African trypanosomiasis in the DRC. PLoS Negl. Trop. Dis. 17, e0011299. (doi:10.1371/journal.pntd.0011299)

35. Mahamat MH et al. 2017 Adding tsetse control to medical activities contributes to decreasing transmission of sleeping sickness in the Mandoul focus (Chad). PLoS Negl. Trop. Dis. 11, e0005792. (doi:10.1371/journal.pntd.0005792)

36. DNDi. 2022 https://dndi.org/wp-content/uploads/2023/10/DNDi-OXA-04-HAT-Clinical-Trial-Protocol-Synopsis.pdf.

