## Supplementary methods and information for "Evolution and impact of the strategy to eliminate *gambiense* human African trypanosomiasis in Guinea"

### S1 Text: Additional methods and information

March 16, 2026

**S1.7 PRIME-NTD criteria** **29**

#### S1.1 New aspects of the modelling compared to previous publications

- This is the first time that we have published a fit of the Warwick gHAT model to epidemiological data from Guinea. Only one other study has modelled the dynamics of gHAT transmission in Guinea (using data for 4 timepoints during 2008–2014) [1] and another has explored the role of possible asymptomatic contribution to transmission using an analysis of the basic reproduction number, $R_0$, with one timepoint of data (in 2007) [2].
- This version of the model has been used to fit to CIV (in prep.) and is based on updates to the models published for the DRC [3], CIV [4], UGA (submitted)and Chad [5].
- Specific to this paper we found that our previously published model of vector dynamics did not fit all the entomological data as well as we would like. In particular, our original model did not capture the sudden increase in tsetse trap catches in Boffa East in 2018. To improve our fits we therefore:

1. changed our fits so that the number of Tiny Targets deployed is directly used as an input and hence our parameter $p_{\text{targetdie}}$ which was the probability of hitting the target and dying immediately after a Tiny Target deployment is now the *increase* in the probability of hitting the target and dying *per target deployed*, and
2. for Boffa East we include an injection of new adult flies in 2018 which are assumed to have migrated from outside the intervention area due to large anthropogenic changes in the area (see main text).

### S1.2 Data

#### S1.2.1 Entomological data

Entomological data started being collected routinely as vector control via Tiny Targets was implemented across the foci. In each region, a pre-deployment monitoring survey to find the average apparent trap density (ATD) was conducted. This was followed by surveys after deployments to monitor how the population changed over time. At each survey in each of Boffa East, Boffa West, and Dubréka, 20 sentinel traps were placed at the same sites so that catch data could be compared. In Forécariah, 31 traps were used.

Boffa East was the first focus to do deployments for Tiny Targets in 2012 and the T0 survey was in May 2011. Since this time monitoring with traps has taken place 2 or 3 times per year.

Vector control started in Dubréka and Boffa West in 2016, with the T0 monitoring surveys in November 2015. Forécariah started VC in 2018 with the T0 survey October 2017. All of these foci have had entomological monitoring (traps) twice per year since then.

All data used for parameterising the interventions (Tiny Targets) and for fitting tsetse reductions (trap data) in this model can be found in the Supplementary Material file `S2-vector_control_data.xlsx`.

#### S1.2.2 Epidemiological data

##### S1.2.2.1 Ebola interruptions

Following information provided in Camara et al. [6] we know that passive screening was reinforced from early 2014 by introducing RDTs in peripheral health facilities (around 30 facilities per focus). Based on the monthly number of people screened passively, there is a sharp improvement from May 2014 (see Figure A). We therefore simulate a rapid improvement in both the stage 1 and stage 2 detection rates in PS from May 2014.

From February 2015 we also see from the same data that there is a marked drop in the number of people tested in PS due to the continuing Ebola outbreak in Guinea. There were no PS tests performed between July and October 2015.

#### S1.2.3 Data extraction into foci

We are primarily interested in four extant foci: Boffa East, Boffa West, Dubréka and Forécariah. Main text Figure 1 shows the boundaries we consider to define each focus.

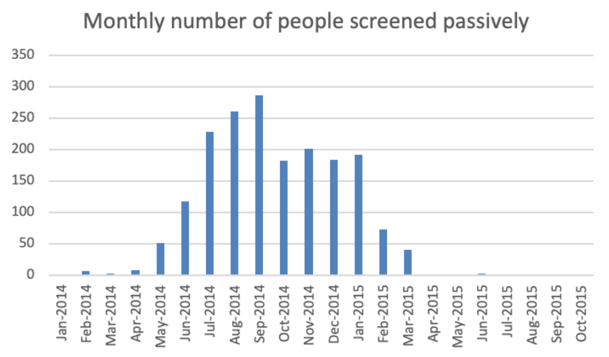

Fig A: Number of people screened passively each month across all foci from January 2014 to October 2015. Data available from Camara et al. [6].

###### S1.2.3.1 WHO HAT Atlas data

One of our sources of epidemiological data was the WHO HAT Atlas spanning the years 2000—2022. It included all reported data from Guinea for this period including the number of active cases, passive cases and number of people actively screened. Some of the data reported are from historical foci and these records were removed from our data set.

We used the focus as recorded in the WHO HAT Atlas as the primary indicator of the disease focus. However, this does not differentiate between Boffa East and Boffa West (which is an informal categorisation based on the different history of VC in the two areas), therefore geolocation was used to allocate records from Boffa to Boffa East and Boffa West, with records allocated to Boffa East or Boffa West if they were recorded as Boffa focus and were either in the Boffa East (West) polygon or Boffa East (West) was the nearest of the two polygons to the record's geolocation.

Having finalised focus allocation, the data were aggregated within focus and year to give counts of:

- the number of people actively screened,
- the number of new active stage 1 cases,
- the number of new active stage 2 cases,
- the number of new active cases with unknown stage,
- the number of new passive stage 1 cases,
- the number of new passive stage 2 cases, and
- the number of new passive cases with unknown stage.

###### S1.2.3.2 PNLMTN-PCC data

For the years 2023–2024 we used the PNLMTN-PCC data for active and passive cases and the number of people actively screened per focus per year. PNLMTN-PCC aggregated these data before sending them.

##### S1.2.4 Population

The population values used in the transmission modelling are at-risk population estimates, the active focus areas being much smaller than the prefecture/subprefecture areas associated with the foci for Boffa West, Dubréka and Forécariah. Baseline estimates of the at-risk population came from intensive population surveys carried out in 2011 [7] in Boffa East and Boffa West and in 2008 in Forécariah[8], and a best-knowledge estimate of 20,000 for Dubréka, assumed to apply to the year in which VC started (2016). These estimates were adjusted to 2024, assuming exponential growth, see Table A.

Population growth rates over the period 1996–2014[9] were used in the adjustment of the at-risk population estimates to 2024 and in the modelling assuming exponential growth. These are presented in Table A. The values for Dubréka and Forécariah are at the prefectural level, whereas the values for the Boffa East and Boffa West foci were calculated from the combination of subprefectures that define them.

Table A: At-risk population estimates and population growth rates.

| Focus | 2024 at-risk population | population growth rate |
|---|---|---|
| Boffa East | 20,464 | 1.019 |
| Boffa West | 5,639 | 1.018 |
| Dubréka | 30,694 | 1.055 |
| Forécariah | 22,132 | 1.013 |

#### S1.3 The compartmental gHAT model

The gHAT models we considered in this study were variants 1–9 of the Warwick model previously presented in the literature [5, 10–14] and are summarised in Table B. gHAT infections among hosts are described by equation (S1.3.1) for the models including animal transmission. The models without animal transmission

Table B: Different model structures considered

| Model | Participation in active screening | | | | Cryptic transmission | |
|---|---|---|---|---|---|---|
| | Random | | None | | Animals | Asymptomatic human infections |
| | gHAT infection risk | | gHAT infection risk | | | |
| | Low | High | Low | High | | |
| M1 | ✓ | | | | | |
| M2 | ✓ | ✓ | | | | |
| M3 | ✓ | | ✓ | | | |
| M4 | ✓ | | | ✓ | | |
| M5 | ✓ | ✓ | ✓ | ✓ | | |
| M6 | ✓ | | | | ✓ | |
| M7 | ✓ | | | ✓ | ✓ | |
| M8 | ✓ | ✓ | ✓ | ✓ | ✓ | |
| M9 | ✓ | | | ✓ | | ✓ |

use the same equations for human hosts and vectors, but not the animal host equations. Human hosts are modelled by a susceptible-exposed-infectious-infectious-recovered-susceptible (SEIIRS) model with two infectious compartments, stage 1 disease, $I_{1H}$, and stage 2 disease, $I_{2H}$. Animal hosts that contribute to transmission are described by a susceptible-exposed-infectious model, with infected animals remaining infectious for life.

Vectors are modelled by using compartments to appropriately model tsetse when used in a host-vector model with disease [12]. Pupal stage tsetse, $P_V$, emerge into unfed susceptible adults, $S_V$, and following a blood-meal become either exposed, $E_V$, or have reduced susceptibility to the *Trypanosoma brucei gambiense* parasites, $G_V$ - this effect is known as the teneral phenomenon. Following an infection, tsetse have an extrinsic incubation period (EIP) before becoming onwardly infectious. To incorporate a more realistic EIP distribution, there are three exposed classes, $E_{1V}, E_{2V}, E_{3V}$, which results in a gamma-distributed EIP (rather than an exponential EIP with a single exposed class).

In order to reduce the dimensionality of our ODE system (by one), the vector equations are non-dimensionalised using the scaling $N_H/N_V$, where $N_H$ is the total human population, and $N_V$ is the tsetse population size. This results in a new non-dimensionalised parameter, $m_{\text{eff}}$, which is $\frac{p_H N_V}{N_H}$ appearing in host equations ($p_H$ is the probability of a human being infected by a single infectious bloodmeal) and is referred to as the *effective vector density*.

The human population in each model variant is partitioned into up to four groups based on combinations of risk of infection (low or high) and participation in active screening (none or at random), see Table B. The proportion of tsetse bites taken on the different groups of humans are $f_1, f_2, f_3 f_4$, depending on the relative availability/attractiveness and the relative abundance of the different groups. High-risk humans are assumed to be $r$-fold more likely to receive bites, i.e. $s_1 = 1$ and $s_4 = r$. Therefore, $f_i$'s can be calculated using $f_i = \frac{s_i N_{Hi}}{\sum_j s_j N_{Hj}}$.

$$
\text{Humans}\;\left\{
\begin{aligned}
\frac{\mathrm{d}S_{Hi}}{\mathrm{d}t} &= \mu_H N_{Hi} + \omega_H^s I_{1Hi}^s + \omega_H^b I_{1Hi}^b + \omega_H R_{Hi} - \alpha m_{\text{eff}} f_i \frac{S_{Hi}}{N_{Hi}} I_V - \mu_H S_{Hi} \\
\frac{\mathrm{d}E_{Hi}}{\mathrm{d}t} &= \alpha m_{\text{eff}} f_i \frac{S_{Hi}}{N_{Hi}} I_V - (\sigma_H + \mu_H) E_{Hi} \\
\frac{\mathrm{d}I_{1Hi}^s}{\mathrm{d}t} &= \sigma_H (1 - p_{bs}) E_{Hi} - (\theta + \omega_H^s + \mu_H) I_{1Hi}^s \\
\frac{\mathrm{d}I_{1Hi}^b}{\mathrm{d}t} &= \sigma_H p_{bs} E_{Hi} + \theta I_{1Hi}^s - (\varphi_H + \omega_H^b + \eta_H(Y) + \mu_H) I_{1Hi}^b \\
\frac{\mathrm{d}I_{2Hi}}{\mathrm{d}t} &= \varphi_H I_{1Hi}^b - (\gamma_H(Y) + \mu_H) I_{2Hi} \\
\frac{\mathrm{d}R_{Hi}}{\mathrm{d}t} &= \eta_H(Y) I_{1Hi}^b + \gamma_H(Y) I_{2Hi} - (\omega_H + \mu_H) R_{Hi}
\end{aligned}
\right.
$$

$$
\text{Animals}\;\left\{
\begin{aligned}
\frac{\mathrm{d}S_A}{\mathrm{d}t} &= \mu_A N_A - \alpha m_{\text{eff}} f_A \frac{S_A}{N_A} I_V - \mu_A S_A \\
\frac{\mathrm{d}E_A}{\mathrm{d}t} &= \alpha m_{\text{eff}} f_A \frac{S_A}{N_A} I_V - (\sigma_A + \mu_A) E_A \\
\frac{\mathrm{d}I_A}{\mathrm{d}t} &= \sigma_A E_A - \mu_A I_A
\end{aligned}
\right.
$$

$$
\text{Tsetse}\;\left\{
\begin{aligned}
\frac{\mathrm{d}P_V}{\mathrm{d}t} &= B_V N_H - (\xi_V + \tfrac{P_V}{K}) P_V \\
\frac{\mathrm{d}S_V}{\mathrm{d}t} &= \xi_V \mathbb{P}(\text{pupating}) P_V - \alpha S_V - \mu_V S_V \\
\frac{\mathrm{d}E_{1V}}{\mathrm{d}t} &= \alpha\big(1 - f_T(t)\big) p_V \big(\textstyle\sum_i f_i \dfrac{(I_{1Hi}^b + x I_{1Hi}^s + I_{2Hi})}{N_{Hi}} + f_A \dfrac{I_A}{N_A}\big)(S_V + \varepsilon G_V) \\
&\qquad -(3\sigma_V + \mu_V + \alpha f_T(t)) E_{1V} \\
\frac{\mathrm{d}E_{2V}}{\mathrm{d}t} &= 3\sigma_V E_{1V} - (3\sigma_V + \mu_V + \alpha f_T(t)) E_{2V} \\
\frac{\mathrm{d}E_{3V}}{\mathrm{d}t} &= 3\sigma_V E_{2V} - (3\sigma_V + \mu_V + \alpha f_T(t)) E_{3V} \\
\frac{\mathrm{d}I_V}{\mathrm{d}t} &= 3\sigma_V E_{3V} - (\mu_V + \alpha f_T(t)) I_V \\
\frac{\mathrm{d}G_V}{\mathrm{d}t} &= \alpha(1 - f_T(t))\Big(1 - p_V \big(\textstyle\sum_i f_i \dfrac{(I_{1Hi}^b + x I_{1Hi}^s + I_{2Hi})}{N_{Hi}} + f_A \dfrac{I_A}{N_A}\big)\Big) S_V \\
&\qquad -\alpha\Big(f_T(t) + \big(1 - f_T(t)\big) p_V \varepsilon \big(\textstyle\sum_i f_i \dfrac{(I_{1Hi}^b + x I_{1Hi}^s + I_{2Hi})}{N_{Hi}} + f_A \dfrac{I_A}{N_A}\big)\Big) G_V \\
&\qquad -\mu_V G_V
\end{aligned}
\right.
\tag{S1.3.1}
$$

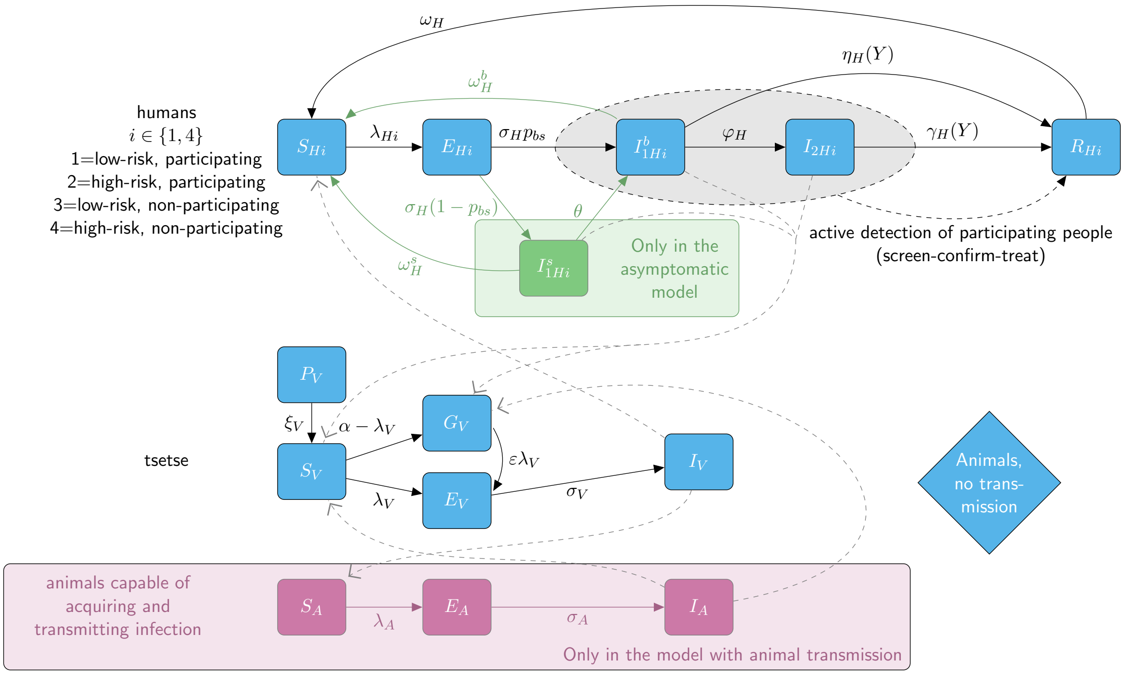

Fig B: Compartmental gHAT model. Unlike the main text version which shows descriptions of the boxes, this version shows the boxes labelled in accordance with Equations S1.3.1 and also shows the rates of transistion between compartments as greek letters next to the corresponding arrows.

##### S1.3.1 Model parameterisation

###### S1.3.1.1 Over-dispersion parameters

Observed numbers of cases, from both active and passive screening, are assumed to be Beta-Binomially distributed, $\mathrm{BetBin}(n, \alpha, \beta)$ where the probability $p$ associated with the binomial distribution is not fixed but a random draw from a Beta distribution with parameters $\alpha$ and $\beta$. The mean ($\mu$) and variance ($\sigma^2$) of this distribution are:

$$\mu = \frac{n\alpha}{\alpha + \beta} \tag{S1.3.2}$$

$$\sigma^2 = \frac{n\alpha\beta(\alpha + \beta + n)}{(\alpha + \beta)^2(\alpha + \beta + 1)} = np(1 - p)\frac{\alpha + \beta + n}{\alpha + \beta + 1} = np\,(1 - p)\,[1 + (n - 1)\,\rho] = np\,(1 - p)\,\kappa \tag{S1.3.3}$$

From this, $\kappa = 1 + (n - 1)\,\rho$ and $\rho = \frac{\kappa - 1}{n - 1}$ allowing us to choose between estimating either $\rho$ or $\kappa$ within the model fitting and easily flip between parameterizations. We have previously used and fitted the intra-class correlations, $\rho_{\text{act}}$ and $\rho_{\text{pass}}$, with priors $\rho_{\text{act}} \sim \mathrm{Beta}(1, 2499)$ and $\rho_{\text{pass}} \sim \mathrm{Beta}(1, 35713)$, respectively, at the health zone level in DRC[15]. Here, we fit $\kappa_{\text{act}}$ and $\kappa_{\text{pass}}$ with priors as given in Table D.

Table C: **Model parameterisation (fixed parameters).** Notation, a brief description, and the values used for fixed parameters.

| Notation | Description | Value | |
|---|---|---|---|
| $N_H$ | Total human population size in 2024 | Varies by focus | Table A |
| $\mu_H$ | Natural human mortality rate | $4.827 \times 10^{-5}$ days$^{-1}$ | [16]* |
| $B_H$ | Total human birth rate | $= \mu_H N_H$ | |
| $\sigma_H$ | Human incubation rate | 0.0833 days$^{-1}$ | [17] |
| $\varphi_H$ | Stage 1 to 2 progression rate | 0.0019 days$^{-1}$ | [18, 19] |
| $\omega_H$ | Recovery rate or waning-immunity rate | 0.1 days$^{-1}$ | [20] |
| Sens(AS) | Active screening algorithm diagnostic sensitivity | 0.766 | ** |
| Spec(AS) | Active screening algorithm diagnostic specificity | 1 | ‖ |
| $B_V$ | Tsetse birth rate (per capita rate of depositing new pupae) | Varies by focus | Table F |
| $f_{\text{max}}$ | Per target and per blood meal probability of a tsetse contacting a Tiny Target and dying | Varies by focus | Table F |
| $\xi_V$ | Rate of pupal development to adult flies | 0.037 days $^{-1}$ | [21, 22] |
| $\mathbb{P}$(pupating) | Probability of a pupa surviving to emerge as an adult fly | 0.75 | [22]‡ |
| $\mu_V$ | Tsetse mortality rate | 0.03 days$^{-1}$ | [21] |
| $\sigma_V$ | Tsetse incubation rate | 0.034 days$^{-1}$ | [23, 24] |
| $\alpha$ | Tsetse bite rate | 0.333 days$^{-1}$ | [21, 25] |
| $p_V$ | Probability of tsetse infection per single infective bite | 0.29 | [26] |
| $\varepsilon$ | Reduced susceptibility factor for non-teneral (previously fed) flies | 0.17 | [26]§ |
| $f_H$ | Proportion of blood-meals on humans by *G. palpalis* | 0.18 | [27] |
| $d_{\text{change}}$ | Midpoint year for passive improvement | 2014.33 | [6] (see Section S1.3.2) |
| $d_{\text{steep}}$ | Speed of improvement in passive detection rate | 100 | |
| **Parameters specific to the models with animal transmission (M6–M8)¶...** | | | |
| $\mu_A$ | Natural animal mortality rate | 0.0014 days$^{-1}$ | Assumed |
| $\sigma_A$ | Animal incubation rate | 0.0833 days$^{-1}$ | [17] |

*The value is the reciprocal of the life expectancy at birth for 2010 (56.72 years).
**Expert opinion, taking into consideration published results [28–47].
‖Source: PNLMTN, due to parasitological confirmation.
‡Pupal survival computed based on a 1% per day mortality rate of pupae over 27 days.
§Whilst the teneral phenomenon is well known, the exact value for the reduction in susceptibility is unknown and likely depends on the age and nutritional status of fed flies [48]. Previous modelling demonstrates that this parameter is non-identifiable in model fitting and will be highly correlated with $R_0^2$ which is fitted [11].
¶M6–M8 as defined in table B.

Table D: **Model parameterisation (fitted parameters).** Notation, relevant models, brief description, and information on the prior distributions for fitted parameters.

| Notation | Model(s)* | Description | Prior distribution† | Percentiles of prior distribution [2.5, 50 & 97.5%] | Unit |
|---|---|---|---|---|---|
| $R_0$ | M1–M9 | Basic reproduction number (NGM approach) | $1+\text{Exp}(10)$ | [1.003, 1.069, 1.369] | - |
| $r$ | M2,M4,M5, M7–M9 | Relative bites taken on high-risk humans | $1+\Gamma(3.68, 1.09)$ | [2.015, 4.654, 10.028] | - |
| $\mathbf{k}=[k_1,k_2]$ | M2,M5,M8 | Human population classificiation‡ | $\text{Dir}(16.97, 3.23)$ | $k_1:[0.656, 0.851, 0.961]$<br>$k_2:[0.039, 0.149, 0.344]$§ | - |
| $\mathbf{k}=[k_1,k_3]$ | M3 | Human population classificiation‡ | $\text{Dir}(16.97, 3.23)$ | $k_1:[0.656, 0.851, 0.961]$<br>$k_3:[0.039, 0.149, 0.344]$§ | - |
| $\mathbf{k}=[k_1,k_4]$ | M4,M7,M9 | Human population classificiation‡ | $\text{Dir}(16.97, 3.23)$ | $k_1:[0.656, 0.851, 0.961]$<br>$k_4:[0.039, 0.149, 0.344]$§ | - |
| $\mathbf{k}=[k_1,\ldots,k_4]$ | M5,M8 | Human population classificiation‡ | $\text{Dir}(16.97, 1.08, 1.08, 1.08)$ | $k_1:[0.656, 0.851, 0.961]$<br>$k_{j,j\neq 1}:[0.002, 0.039, 0.182]$§ | - |
| $\eta_H^{\text{post}}$ | M1–M9 | Treatment rate from stage 1, 1998 onwards | $\Gamma(3.54, 5.32\times 10^{-5})$ | [4.59, 17.1, 42.9]$\times 10^{-5}$<br>[7.12, 20.7, 45.7]$\times 10^{-5}$ | days$^{-1}$ |
| $\gamma_H^{\text{post}}$ | M1–M9 | Combined treatment and disease-induced death rate from stage 2, 1998 onwards | $\Gamma(6.21, 0.001)$ | [0.00233, 0.00588, 0.0120] | days$^{-1}$ |
| $\gamma_H^{\text{pre}}$ | M1–M9 | Relative treatment/death rate from stage 2 factor, pre-1998 | $\Gamma(6.21, 0.001)$ | [0.00233, 0.00588, 0.0120] | - |
| $u$ | M1–M9 | Proportion of stage 2 passive cases reported | $\text{B}(20, 40)$ | [0.2208, 0.3315, 0.4564] | - |
| $\eta_{H_{\text{amp}}}$ | M1–M9 | Relative improvement in passive stage 1 detection rate | $\Gamma(2.17, 0.85)$ | [0.253, 1.57, 4.98] | - |
| $\gamma_{H_{\text{amp}}}$ | M1–M9 | Relative improvement in passive stage 2 detection rate | $\Gamma(2.17, 0.85)$ | [0.253, 1.57, 4.98] | - |
| $\kappa_{\text{act}}$ | M1–M9 | Overdispersion parameter for active detection | $1+\Gamma(1, 0.8)$ | $[1.02, 1.55, 3.95]$ | - |
| $\kappa_{\text{pass}}$ | M1–M9 | Overdispersion parameter for passive detection | $1+\Gamma(1, 0.28)$ | $[1.01, 1.19, 2.03]$ | - |
| $f_A$ | M6–M8 | Proportion of blood meals on animals able to acquire and transmit the parasite | $\text{B}(1.3, 1.3)$ | [0.046, 0.500, 0.954] | - |
| $k_A$ | M6–M8 | Relative size of the animal population able to acquire and transmit the parasite | $\Gamma(1.26, 19.3)$ | [1.18, 18.3, 81.4] | - |
| $p_{bs}$ | M9 | Proportion of human exposures resulting in initial blood infection | $\text{B}(10, 4)$ | [0.462, 0.725, 0.909] | - |
| $\omega_H^b$ | M9 | Self-cure rate for stage 1 blood infections | $\Gamma(4, 1.5\times 10^{-6})$ | [0.163, 0.551, 1.32] $\times 10^{-5}$ | days$^{-1}$ |
| $\omega_H^s$ | M9 | Self-cure rate for skin-only infections | $\Gamma(4, 10^{-4})$ | [1.09, 3.67, 8.77] $\times 10^{-4}$ | days$^{-1}$ |
| $\theta$ | M9 | Transition rate from skin-only to blood infection | $\Gamma(4, 10^{-5})$ | [1.09, 3.67, 8.77]$\times 10^{-5}$ | days$^{-1}$ |
| $x$ | M9 | Relative infectiousness of skin-only infection compared to a blood infection | $\text{B}(3, 3)$ | [0.147, 0.500, 0.853] | days$^{-1}$ |

*Models as defined in Table B.
†Where Exp(.), Γ(.), B(.) and Dir(.) are the exponential, gamma (parameterised with shape and scale), beta and Dirichlet distributions, respectively.
‡Human population classification: $k_1$ is the proportion of humans at low risk of infection that participate at random in AS; $k_2$ is the proportion of high-risk humans participating at random in AS; $k_3$ is the proportion of low-risk humans that do not participate in AS, and $k_4$ is the proportion of high-risk humans that do not participate in AS.
§Percentiles of marginal (beta) distributions.

##### S1.3.2 Modelling passive detection and its improvement

In this study we follow similar methods for modelling passive detection and its improvement as Crump et al. [10] and this is described below for completeness. The key change is how we deal with interruptions in PS due to the Ebola outbreak.

There are two sources of passive detection improvements considered in our model: a rapid improvement due to the introduction of the card agglutination test for trypanosomes (CATT) test in all foci in 1998 and a second improvement due to the introduction of rapid diagnostic tests (RDTs) in 2014. We also simulated temporary PS worsening in 2015 due to Ebola. Prior distributions and percentiles of parameters related to passive detection and its improvement and worsening over time are summarised in Table D. Fixed values were used for both $d_{\text{change}}$ and $d_{\text{steep}}$ because the passive screening was reinforced rapidly at the beginning of the Ebola outbreak, these values are given in Table C.

For improvements from 1998 we use the following equations to describe transmission rates from infected classes:

$$\eta_H(Y) = \eta_H^{\text{post}} \left[ 1 + \frac{\eta_{H_{\text{amp}}}}{1 + \exp\left(-d_{\text{steep}}(Y - d_{\text{change}})\right)} \right] \tag{S1.3.4}$$

$$\gamma_H(Y) = \gamma_H^{\text{post}} \left[ 1 + \frac{\gamma_{H_{\text{amp}}}}{1 + \exp\left(-d_{\text{steep}}(Y - d_{\text{change}})\right)} \right] \tag{S1.3.5}$$

We assume that all stage 1 cases are reported, but that some of the exits from stage 2 are due to death from gHAT disease. In 1998 the reporting probability for an exit from stage 2 is given by $u$, however as the exit rate from stage 2 increases this reporting probability does not stay constant, but increases (proportionally more people would be detected and treated with higher exit rates). When we compute reporting rates from stage 2 we therefore use the following:

$$\text{Death rate} = (1 - u)\gamma_H^{\text{post}} \tag{S1.3.6}$$

$$\text{Stage 2 reporting incidence} = (\gamma_H(Y) - \text{Death rate})(I_{2H1} + I_{2H4}) \tag{S1.3.7}$$

Improvement in passive screening in all foci is assumed from May 2014 due to increased testing (see Section S1.2.2.1). The functional form of PS improvement in the model is a simple sigmoidal model which has been used in other countries (e.g. DRC [3, 10] and Chad [5]) and is able to capture gradual improvements over time or sudden improvements depending on its parameterisation.

For Guinea we have evidence from the PS testing numbers that there was a rapid change in May 2014, therefore we set$d_{\text{change}} = 2014.X$ and $d_{\text{steep}} = 100$. What is less clear is the rate of detection for stage 1 and stage 2 from the point of infection before 2014 and after so these rates were estimated for each focus separately in our model fitting procedure.

Prior distributions for the PS fitted parameters can be found in Table D.

For the worsening of PS in 2015, we assumed there was a sudden reduction in detection rate back to the pre-May 2014 values in February 2015, and that testing in PS stopped completely in July 2015 for the remainder of the year. From January 2016 we assumed PS detection rates jumped back to the same values as the beginning of 2015.

##### S1.3.3 Modelling vector control

In the present study we utilise an evolution of the method to simulate the impact of annual vector control on tsetse populations presented elsewhere including for location with twice yearly deployments [4, 10, 12, 49]. We model the dynamics of tsetse populations in the presence of Tiny Target-based vector control based on an adjustment to historical tsetse density data for the four foci. In general Tiny Targets have been found to be highly effective (typically at least 80% density reduction after one year) across different settings [7, 50–52]. Unlike in previous fitting, this time we adjust for the number of tiny targets deployed in a given deployment.

The function which describes the probability of a host-seeking tsetse both hitting a Tiny Target and dying as a result, $f_T$, is time-dependent ($t$, the time in days since the targets were most recently deployed). It also depends on $n_{\text{target}}$, the number of targets deployed in the most recent deployment and $f_{\text{max}}$ - the maximum daily probability of contacting a Tiny Target and dying as a result *per target*:

$$f_T(t) = f_{\text{max}} n_{\text{target}} \left(1 - \frac{1}{1 + \exp\left(-VC_{\text{steep}}(t - VC_{\text{change}})\right)}\right) \qquad \text{(S1.3.8)}$$

N.B. In previous work by our group $f_{\text{max}}$ was used to describe the total probability of hitting a target and dying across all deployed targets and so not multiplied by the number of targets. This new formulation is more flexible as it allows the model to use a variable number of Tiny Targets deployed in different years and create a corresponding change in the modelled tsetse dynamics.

We assume that the targets are set out over a sufficiently large area and sufficiently sparsely compared to typical tsetse movements that the probability of a fly contacting with a target and dying is linear in the number of targets. $f_T$ modifies all the bite rates $\alpha$ in our tsetse equations to produce an additional Tiny-Target-induced mortality for tsetse. The effectiveness of Tiny Targets is assumed to wane over time so that at the point of the next deployment, the previous deployment is virtually non-effective. To simulate this we have a rapid decrease in effectiveness after $VC_{\text{change}}$ days. This represents lost Tiny Targets (e.g. due to rainfall) or loss of the effectiveness of remaining targets (e.g. vegetation growth impacting their visibility).

In Boffa East, we noted that there was a strong resurgence in the fly density data from 2018 onwards. The model as specified would not be able to capture this, so to allow this to be captured, we added an instantaneous tsetse reintroduction in 2018. In reality, we believe the issue is less that the fly population grew back quickly, more that the movement of animals caused by human pressure brought the flies with them from neighbouring areas that weren't reached by the vector control, but that adding more flies is a good approximation of this reinvasion. The reinvasion was assumed to be equivalent to adding some proportion of the pre-intervention equilibrium population of susceptible adult flies (teneral and non-teneral) to the state. The size of this reinvasion was fitted as a parameter. Due to lack of any specific information on when this reinvasion would occur, we assumed it would happen at the date of the first data point after the resurgence, which was taken to be at the start of May 2018.

#### S1.4 Fitting to data

##### S1.4.1 Fitting to entomological data

The tsetse model is adjusted to entomological data from each focus separately using a maximum likelihood estimation on the parameters $f_{\text{max}}$ and $B_V$. In Boffa East we also estimate the size of the re-invasion of adult tsetse $\text{Reinvasion}_{\text{size}}$. The parameters are optimised using the Differential Evolution evolutionary algorithm [53], specifically the R function `DEoptimR::JDEoptim`[54, 55]. The fitting was replicated 10 times in each focus, starting from randomly generated populations of possible parameters in the ranges specified in Table E. The best solutions from each of the 10 replicates are summarised in Table F.

$$\begin{aligned} LL(\theta, g(0)|\text{VC data}) &= \log(\mathbb{P}(\text{VC data}|\theta, g(t_0))) \\ &= \log\left(\prod_i \mathbb{P}(n_i = g(t_i)|\theta, g(t_0))\right) \\ &= \sum_i \log\left(\mathbb{P}(n_i = g(t_i)|\theta, g(t_0))\right) \\ &= \sum_i \log\left(\frac{g(t_i)^{n_i} e^{-g(t_i)}}{n_i!}\right) \\ &= \sum_i \left(n_i \log g(t_i) - g(t_i) - \log n_i!\right) \end{aligned} \qquad \text{(S1.4.1)}$$

The best model fits to the catch data, $g(t)$, and functions $f_T(t)$ for each focus are shown in Figure 3 in the main paper. Fitted parameter values for each focus are reported in Table F.

Table E: The parameters for fitting the vector population model

| **Parameter** | **Description** | **Fitted** | **Value** Minimum | Maximum |
|---|---|---|---|---|
| scale | Scale factor for matching $f_t$ to catch totals | Fitted | 1 | 600 |
| $p_{\text{TargetDie}}$ | Maximum efficacy of tiny targets (per target) | Fitted | 0 | 1 |
| $B_V$ | Birth rate | Fitted | $\frac{\mu_V}{p_{\text{survive}}}$ | $\frac{365}{2}$ |
| $\text{Reinvasion}_{\text{size}}$ | Level of re-invasion as a proportion of pre-intervention adult tsetse population size (Boffa East only) | Fitted | 0 | 1 |
| $\text{Reinvasion}_{\text{year}}$ | Time of re-invasion (Boffa East only) | Fixed | 1 May 2018 | |
| $VC_{\text{steep}}$ | Steepness of the decline in tiny target effectiveness | Fixed | $25$ days$^{-1}$ | |
| $VC_{\text{change}}$ | Days from deployment until tiny targets hit 50% effectiveness | Fixed | $0.35 \cdot 365$ days | |
| $\xi_V$ | Death rate | Fixed | $0.0370$ days$^{-1}$ | |
| $p_{\text{survive}}$ | Probability that a pupa survives the pupal stage to become an adult | Fixed | 0.75 | |
| $\alpha$ | Tsetse biting rate | Fixed | $0.333$ days$^{-1}$ | |
| $\mu_V$ | Death rate | Fixed | $0.03$ days$^{-1}$ | |

###### S1.4.1.1 Results

Table F: Summary of tsetse population model parameters across ten replicated model fits.

| Focus | Value | Parameter | | | | -LogLik |
|---|---|---|---|---|---|---|
| | | scale | $B_V$ | $f_{\text{max}}$ | $\text{Reinvasion}_{\text{size}}$ | |
| Boffa East | Min | 116.9615 | 19.0990 | $1.1294\times10^{-5}$ | 0.5451 | 346.2004 |
| | Max | 117.0919 | 19.1106 | $1.1326\times10^{-5}$ | 0.5458 | 346.2010 |
| | Best | 117.0919 | 19.1106 | $1.1326\times10^{-5}$ | 0.5452 | 346.2004 |
| Boffa West | Min | 278.9017 | 19.7451 | $1.9519\times10^{-5}$ | NA | 141.8717 |
| | Max | 279.5693 | 19.7637 | $1.9594\times10^{-5}$ | NA | 141.8721 |
| | Best | 279.1373 | 19.7530 | $1.9550\times10^{-5}$ | NA | 141.8717 |
| Dubréka | Min | 135.5528 | 26.7881 | $3.3821\times10^{-5}$ | NA | 229.6909 |
| | Max | 135.6513 | 26.8162 | $3.3852\times10^{-5}$ | NA | 229.6915 |
| | Best | 135.6117 | 26.8162 | $3.3852\times10^{-5}$ | NA | 229.6909 |
| Forécariah | Min | 446.0153 | 21.0200 | $1.7950\times10^{-5}$ | NA | 177.4143 |
| | Max | 446.3007 | 21.0303 | $1.7970\times10^{-5}$ | NA | 177.4148 |
| | Best | 446.1854 | 21.0282 | $1.7968\times10^{-5}$ | NA | 177.4143 |

##### S1.4.2 Fitting to epidemiological data

###### S1.4.2.1 Running the models

Our ODE models were run assuming that, prior to 1998, they were at their endemic equilibrium. This endemic equilibrium was calculated analytically, based on model parameterisation, and used as an initial condition in the model code. In 1998 we assume active screening began at the same level reported in 2000 and that there was improvement to passive screening due to the availability of the CATT diagnostic test. In our model this has the effect of perturbing the dynamics away from endemic equilibrium and reducing transmission.

For fitting the models, there are two elements, each of which is initialised.

- Two chains are run in the Markov chain Monte Carlo (MCMC) used in the fitting. The chains are initialised using the fixed parameters and by random perturbations around supplied, individually valid, initial values of each parameter being fitted, rejecting those parameter sets that do not produce a valid posterior probability.
- Each projection is initialised with a randomly sampled realisation from the posterior distribution of fitted parameters alongside the set of fixed parameters.

###### S1.4.2.2 Likelihood

Eight parameters; $R_0, r, \eta_H, \gamma_H, b_{\gamma_{H0}}, k_1, u$, and Spec were fitted in all health zones for both models. A further two parameters; $k_A$ and $f_A$, were fitted in all health zones for the model with animal transmission. Additional parameters were included as required (combinations of $d_{\text{change}}$, $\eta_{H_{\text{amp}}}$, $\gamma_{H_{\text{amp}}}$, $d_{\text{steep}}$ and $b_{\text{specificity}}$ as appropriate, see above).

For fitting the model to case data we transform model ODE solutions (for S1.2.1) into annual case reporting denoted $A_{M1}$, $A_{M2}$, for active stage 1 and stage 2 and $P_{M1}$, $P_{M2}$, for passive stage 1 and 2. Since we always know the stage (1 or 2) in the model simulations there is no requirement for a "U" (unknown stage) category for the model. These are computed using solutions to the ODEs for the given set of parameters aggregated across a year.

In terms of main text Fig 1, they relate to the transfer from infectious categories to the recovered category – the new annual reported case incidence. This is either by passive detection from stage 1 for year $Y$:

$$P_{M1}(Y) = \int_{Y}^{Y+1} \eta_H(Y)\big(I_{1H1}(t) + I_{1H4}(t)\big)\,\mathrm{d}t,$$

passive detection from stage 2

$$P_{M2}(Y) = \int_{Y}^{Y+1} (\gamma_H(Y) - \text{Death rate})\big((I_{2H1}(t) + I_{2H4}(t)\big)\,\mathrm{d}t,$$

or by active screening from the low-risk ($H1$) group in year $Y$

$$A_{M1}(Y) = z(Y)\text{Sens}I_{1H1}(Y) + z(Y)(1 - \text{Spec})\big(k_1 N_H - I_{1H1}(Y) - I_{2H1}(Y)\big)$$

and

$$A_{M2}(Y) = z(Y) \times \text{Sens} \times I_{2H1}(Y)$$

with variable active screening coverage by year, $z(Y)$ and fixed diagnostic sensitivity. $A_{M1}$ also contains any false positives that may have been incorrectly identified from non-infected people based on the high but imperfect specificity of the active screening algorithm. We assume in the DRC that all false positives would be assigned to be stage 1 and treated, however in the model false positives stay in the susceptible class unlike true cases which move to recovereds.

The log-likelihood function used in the adaptive Metropolis-Hastings MCMC contained two terms in each year for which reported case numbers were available for each source of reported cases (active or passive screening). These were:

- a beta-binomial probability that the total number of cases reported in that year for that source came from the available population (either the reported number of people actively screened for active screening, or the health zone population for passive screening) with probability calculated from solving the ODE for the current set of parameters, and

- a binomial probability that the reported stage 1 cases come from the total number of reported staged cases where the probability parameter again comes from the solution of the ODE. In many years staging is unknown and so this part of the log-likelihood will return zero and not contribute to our calculation. In some years, we only partially know staging information.

This formulation allowed over-dispersion in the observed cases to be included, via the beta-binomial distribution, and any proportion of cases with reported disease stage to be appropriately accounted for (assuming that the reporting of staging information is independent of the disease stage). The log-likelihood function was as follows:

$$\begin{aligned} LL(\theta|x) = & \log(P(x|\theta)) \\ \propto & \sum_{i=2000}^{2016} \Bigg( \log\left[\mathrm{BetaBin}\left(A_{D1}(i) + A_{D2}(i) + A_{DU}(i); z(i), \frac{A_{M1}(i) + A_{M2}(i)}{z(i)}, \mathrm{disp_{act}}\right)\right] \\ & + \log\left[\mathrm{Bin}\left(A_{D1}(i); A_{D1}(i) + A_{D2}(i), \frac{A_{M1}(i)}{A_{M1}(i) + A_{M2}(i)}\right)\right] \\ & + \log\left[\mathrm{BetaBin}\left(P_{D1}(i) + P_{D2}(i) + P_{DU}(i); N_H, \frac{P_{M1}(i) + P_{M2}(i)}{N_H}, \mathrm{disp_{pass}}\right)\right] \\ & + \log\left[\mathrm{Bin}\left(P_{D1}(i); P_{D1}(i) + P_{D2}(i), \frac{P_{M1}(i)}{P_{M1}(i) + P_{M2}(i)}\right)\right]\Bigg) \end{aligned}$$

The model takes parameterisation $\theta$, $x$ is the data, $P_{Dj}(i)$ and $A_{Dj}(i)$ are the number of cases detected by passive or active screening (of stage $j$, which may be 1, 2 or unknown, $U$) in year $i$ of the data. $P_{Mj}(i)$ and $A_{Mj}(i)$ are the number of cases detected by passive or active screening (of stage $j$) in year $i$ of the model, and $z(i)$ is the number of people actively screened in year $i$. $\mathrm{BetaBin}(m; n, p, \rho)$ gives the probability of obtaining $m$ successes out of $n$ trials with probability $p$ and overdispersion parameter $\rho$. The overdispersion accounts for larger variance than under the binomial. The probability density function of this distribution is given by:

$$\mathrm{BetaBin}(m; n, p, \rho) = \frac{\Gamma(n+1)\Gamma(m+a)\Gamma(n-m+b)\Gamma(a+b)}{\Gamma(n-m+1)\Gamma(n+a+b)\Gamma(a)\Gamma(b)}$$

where $a = p(1/\rho - 1)$ and $b = a(1-p)/p$.

###### S1.4.2.3 Imputation of missing numbers screened information

There are instances in the data where the number of cases from within year $t$ ($A_D(t) = A_{D1} + A_{D2}$) is not consistent with the number of people recorded as having been screened in that year for that health zone ($z(t)$), i.e. (i) $A_D(t) > z(t)$ or (ii) $A_D(t)/z(t)$ is a much higher prevalence than is biologically realistic for gHAT (e.g. more than 10%).

In the current study, we have chosen to impute the number of negative tests in more situations. We still believe that where the number of cases reported is low these people probably attended screening elsewhere, but imputing a missing screening value, in our independent health zone analyses, is expected to reflect the model's underlying prevalence better. We, therefore, imputed the number of negative tests for a year within a health zone where:

1. the number of cases from was more than 10% of the number screened ($A_D(t) > 0.1 \times z(t)$;
2. the number screened was zero, or not recorded, or less than 20 ($z(t) \leq 20$).

Imputation of the number of negative tests takes place within the MCMC fitting procedure.

We use a Geometric prior for the number of negative screening tests in year $t$, $A_D^-(t) \sim \mathrm{Geom}(\lambda_t)$, where $\lambda_t = \frac{1}{1+\bar{N}_t}$ and $\bar{N}_t = \frac{\sum_{j=2000,j\neq t}^{2020} N_j e^{-|t-j|}}{\sum_{j=2000,j\neq t}^{2020} e^{-|t-j|}}$, a weighted mean of the number of people screened in years other than $t$. The proposal distribution for $A_D^-(t)$ was a negative binomial distribution:

$$A_D^-(t)|A_D(t), p(t) \sim \mathrm{NB}(A_D(t) + 1, 1 - (1 - p(t))\,(1 - \lambda_t))$$

where the probability of active case detection, $p(t)$, was sampled from the following Beta distribution:

$$p(t)|\theta \sim \mathrm{Beta}\left(\hat{p}(t)\left(\frac{1}{\mathrm{disp_{act}}(t)} - 1\right), (1 - \hat{p}(t))\left(\frac{1}{\mathrm{disp_{act}}(t)} - 1\right)\right)$$

and $\hat{p}(t)$ is the probability of active case detection in year $t$ calculated from the ODE outputs.

###### S1.4.2.4 Sequential Bayesian updating for the model with asymptomatic human transmission.

This material is adapted from the Supplementary Information of Crump *et al.*, 2024[14].

There are five parameters which are unique to our model with transmission of HAT to and from a reservoir of asymptomatic human infections:

1. the proportion of human exposures resulting in initial blood infection ($p_{bs}$);
2. the self-cure rate for stage 1 blood infections ($\omega_H^b$);
3. the self-cure rate for skin-only infections ($\omega_H^s$);
4. the transition rate from skin-only to blood infection ($\theta$); and
5. the relative infectiousness of skin-only infection compared to a blood infection ($x$).

We regard these parameters as being innate, biological values and assume that they should be invariant across geographies. This is in contrast to the parameters which are unique to the model with transmission to and from a non-specific animal source, which is liable to vary as a result of differences in the animal species, variability in the amount of animal and tsetse habitats and interactions between these hosts and tsetse.

The best way to fit the asymptomatic model would be to consider all foci together in a hierarchical model, however, this would be more computationally demanding. We have therefore opted to perform Bayesian updating of priors such that posterior parameter distributions from foci that are informative for these parameters provide the prior distributions used for the asymptomatic model-specific parameters in other foci.

The procedure used is as follows:

1. All foci were analysed with the same univariate prior distributions (Table D), five MCMC chains were run in each analysis with 1,000 posterior samples taken from each chain, to give 5,000 posterior samples overall with a minimum effective sample size of 2,500.
2. The Kullback-Liebler divergence of the realised marginal posterior distributions ($Y$) from the univariate, independent prior distributions ($X$), $D_{\mathrm{KL}}(X||Y)$, were calculated and used to assign a rank to the foci $j = 1, \ldots, 4$.
3. Foci $j = 2, \ldots, 4$ were re-analysed using a prior based on the posterior distribution of the five asymptomatic specific parameters from the analysis of focus $j - 1$. A Gaussian mixture model (GMM, a weighted mixture of multivariate normal distributions) was fitted to the samples from the posterior distribution from the analysis of focus $j - 1$.

**GMM fitting** For the MCMC analysis of focus $j$, the 5,000 posterior samples from the analysis of focus $j-1$ were read in. Each of the five asymptomatic model-specific parameters was first transformed to a $(-\infty, +\infty)$ scale and then to have a mean of zero and a standard deviation of one. The transformation to convert to a $(-\infty, +\infty)$ scale and the centring and scaling values are retained to create the Jacobian for the scaled GMM prior.

A Gaussian Mixture Model was fitted to the transformed parameters using the MATLAB routine `fitgmdist`. Akaike's Information Criterion (AIC) was used to select the number of Gaussian distributions to include in the mixture.

A MATLAB structure was created containing the final GMM, identifiers for the parameters, and transformation information and this was saved to act as the multivariate five-parameter prior for the asymptomatic model-specific parameters in the analysis of the $j^{\mathrm{th}}$ focus.

###### S1.4.2.5 Model evidence

We compare our models based on the marginal likelihood, or *evidence*, of the data for each model. The Bayesian model evidence uses the full probability distribution of the model rather than a point estimate (usually the maximum likelihood estimate) and naturally accounts for differences in the number of parameters required for different models.

An importance sampled estimator of the model evidence was implemented following Touloupou et al. [56].

The joint distribution of $(\boldsymbol{\theta}_m, \mathbf{x})$, for parameters $\boldsymbol{\theta}_m = (\theta_1, \theta_2, \ldots, \theta_{d_m})$ of model $m$ and data $\mathbf{x} = (x_1, x_2, \ldots, x_n)$ satisfies

$$\pi(\boldsymbol{\theta}_m|\mathbf{x})\,\pi(\mathbf{x}|\mathbf{m}) = \pi(\mathbf{x}|\boldsymbol{\theta}_m)\,\pi(\boldsymbol{\theta}_m)\,, \tag{S1.4.2}$$

where $\pi(\boldsymbol{\theta}_m|\mathbf{x})$ is the joint posterior distribution of parameters $1 \ldots d$, $\pi(\mathbf{x}|\mathbf{m})$ is the marginal likelihood or *evidence*; $\pi(\mathbf{x}|\boldsymbol{\theta}_m)$ is the likelihood, and $\pi(\boldsymbol{\theta}_m)$ is the prior distribution.

By use of MCMC methods to investigate the posterior distribution of the parameters, calculation of $\pi(\mathbf{x}|\mathbf{m})$ is avoided. Calculation of the evidence for use in model comparison requires computing the integral:

$$\pi(\mathbf{x}|\mathbf{m}) = \int \pi(\mathbf{x}|\boldsymbol{\theta}_m)\,\pi(\boldsymbol{\theta}_m)\,\mathrm{d}\boldsymbol{\theta}_m \tag{S1.4.3}$$

$$= \int \pi(\mathbf{x}|\boldsymbol{\theta}_m)\,\frac{\pi(\boldsymbol{\theta}_m)}{q(\boldsymbol{\theta}_m)} q(\boldsymbol{\theta}_m)\,\mathrm{d}\boldsymbol{\theta}_m \tag{S1.4.4}$$

Equation S1.4.3 cannot be calculated analytically except for some small set of tractable models. It can, however, be rewritten as equation S1.4.4, where $q(\boldsymbol{\theta}_m)$ is a $d_m$-dimensional probability density function. From this, an importance sampled estimator of $\pi(\mathbf{x}|\mathbf{m})$ is:

$$\hat{P}_q = \frac{1}{N}\sum_{i=1}^{N} \pi(\mathbf{x}|\boldsymbol{\theta}_{m,i})\,\frac{\pi(\boldsymbol{\theta}_{m,i})}{q(\boldsymbol{\theta}_{m,i})}, \tag{S1.4.5}$$

where the $\boldsymbol{\theta}_{m,i}$ are $N$ samples drawn from $q$.

A defence mixture [57] was used for $q(\boldsymbol{\theta}_m)$:

$$q(\boldsymbol{\theta}_m) = p\,\phi(\boldsymbol{\theta}^*_m; n, \boldsymbol{\mu}_1 \ldots \boldsymbol{\mu}_n, \mathbf{C}_1 \ldots \mathbf{C}_n) \left|\frac{\boldsymbol{\theta}^*_m}{\boldsymbol{\theta}_m}\right| + (1-p)\,\pi(\boldsymbol{\theta}_m) \tag{S1.4.6}$$

where $\phi(\cdot)$ is a mixture of $n$ multivariate Gaussian distributions with means $\boldsymbol{\mu}_j$ $(j = \{1 \ldots n\})$, and covariance matrices $\mathbf{C}_j$, $\left|\frac{\boldsymbol{\theta}^*_m}{\boldsymbol{\theta}_m}\right|$ is the Jacobian transformation relating probability on transformed and original scales, and $p$ is a mixing proportion ($p = 0.95$ was chosen for use, being a typical value [56]).

In each of our health-zone-level MCMC analyses of the models with and without animal transmission, $2\,000$ samples from the joint posterior distribution were generated and stored, and $\phi(\boldsymbol{\theta}_m; n, \boldsymbol{\mu}_1 \ldots \boldsymbol{\mu}_n, \mathbf{C}_1 \ldots \mathbf{C}_n)$ for each health zone and model was chosen using the Matlab `fitgmdist` function, selecting $n$ based on Akaike's Information Criterion (AIC). To account for the high correlations between some of our model parameters, regularisation was applied to ensure that the covariance matrices, $\mathbf{C}_k$, would be positive semi-definite. Before passing to `fitgmdist`, transformations were applied to the posterior samples to put them in the range $(-\infty, \infty)$ – appropriate for Gaussian distributions – followed by scaling and centring to keep the regularisation consistent across analyses, at least at the simple, single overall covariance matrix level.

Having defined $\phi(\cdot)$ for a given analysis (health zone, model combination), $\hat{P}_q$ was calculated (equation S1.4.5) using $N = 10\,000$ samples drawn from $q(\boldsymbol{\theta}_m)$.

###### S1.4.2.6 Creating an ensemble model

In each focus, $2\,000$ samples from the ensemble model joint posteriors were randomly selected from among the $2\,000$ samples from the joint posterior distribution of the model parameters for each of models 1–8, and the $5\,000$ from the joint posterior samples for model 9 ensemble posterior. The relative evidence, that is the proportion of the total evidence across models from each model, was used as the weight for each model in the ensemble model. Stochastic projections for each focus were performed using the $2\,000$ ensemble model posterior joint samples, with 10 stochastic realisations being created for each parameter set, resulting in $20\,000$ simulations in total.

Fig C: Ensemble model posterior distributions of model parameters.

###### S1.4.2.7 Results

Table G: Relative model evidence (%) – weights used to make up the ensemble model.

| Focus | Model variant | | | | | | | | |
|---|---|---|---|---|---|---|---|---|---|
| | 1 | 2 | 3 | 4 | 5 | 6 | 7 | 8 | 9 |
| Boffa East | <0.01 | <0.01 | <0.01 | 0.02 | 0.01 | 0* | 0.02 | 0* | 99.95 |
| Boffa West | <0.01 | <0.01 | <0.01 | 34.82 | 0.86 | <0.01 | 39.55 | 2.81 | 21.96 |
| Dubréka | <0.01 | <0.01 | <0.01 | 45.59 | 8.33 | 0.01 | 31.40 | 7.11 | 7.55 |
| Forécariah | <0.01 | <0.01 | <0.01 | 27.80 | 1.36 | 0.27 | 33.82 | 0* | 36.75 |

*Model excluded from ensemble due to low effective sample size.

#### S1.5 Counterfactual scenarios

We considered three counterfactual scenarios (CFS). In each CFS, one historical event was assumed not to have taken place so there was either (i) no ebola outbreak, (ii) no improvement to passive screening, or (iii) no implementation of vector control. We simulated each of these CFS and compared them against the actual scenario in which all three events took place (although vector control implementation took place in different years in different foci).

For the modelling of the human population, we utilised a version of the tau-leaping method that employs a hash-based matching pseudo-random number generation technique. This version of the gHAT model is described in Sunnucks et al [58], and ensures that compared realisations of the model 'have the same environmental randomness' by matching the random seed for each event draw at every time step, and replacing the Poisson random variable with a sum of Bernoulli random variables (or a truncated, faster version for the larger compartments). As a result, random events such as when an infected individual progresses from stage 1 to stage 2, tend to occur at the same time across compared realisations, while still allowing for natural stochastic variation. This provides the benefits of the more realistic stochastic modelling, which more accurately captures the dynamics of the disease, without obtaining highly noisy measurements of uncertainty when calculating comparative metrics such as the predicted difference in the number of DALYs between the actual scenario and the CFS.

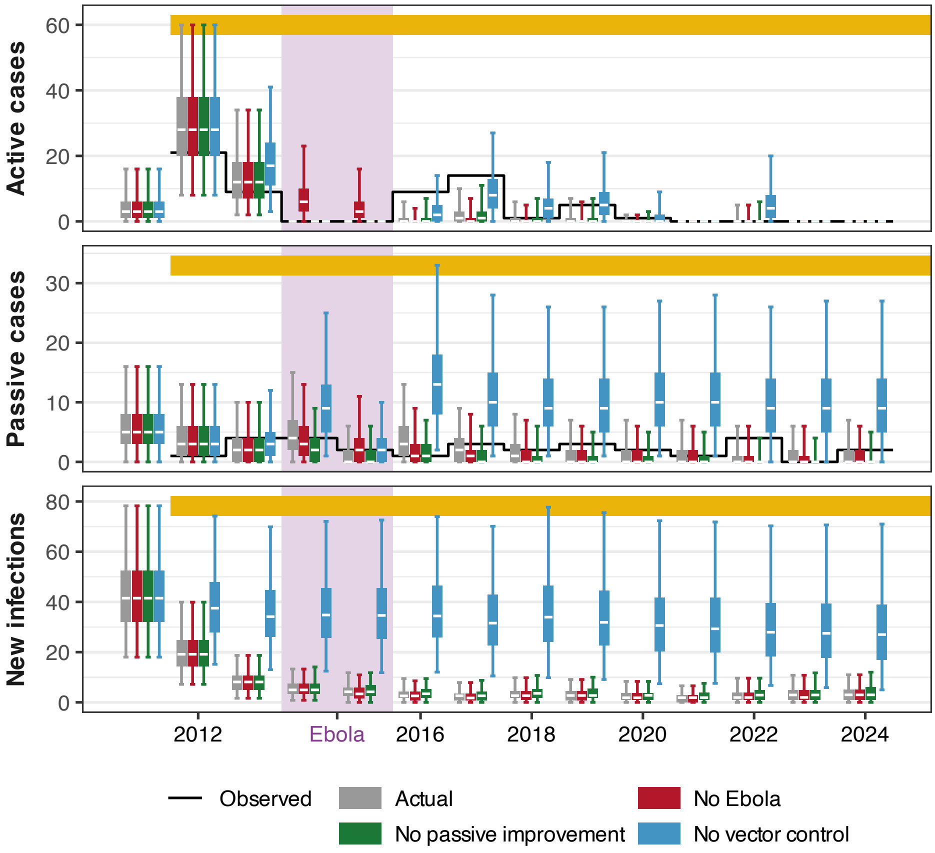

Fig D: Evolution of the number of new reported active and passive cases and new infections each year in Boffa East, according to the actual scenario (grey) and counterfactual scenarios: no Ebola outbreak (red), no improvement in passive screening (green), and no vector control (blue). Boxes and whiskers represent the median and the 50% and 95% prediction intervals. Vector control has been in place in Boffa East since 2012 (yellow bar), and the Ebola outbreak occurred between 2014 and 2016 (light purple background).

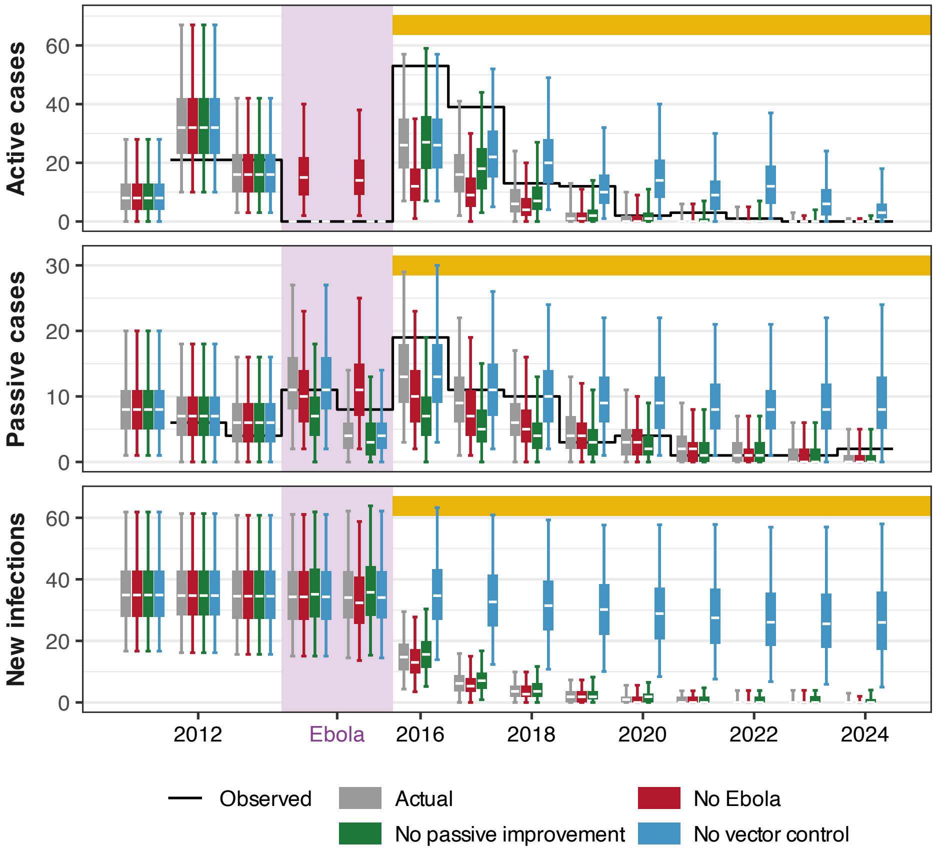

Fig E: Evolution of the number of new reported active and passive cases and new infections each year in Boffa West, according to the actual scenario (grey) and counterfactual scenarios: no Ebola outbreak (red), no improvement in passive screening (green), and no vector control (blue). Boxes and whiskers represent the median and the 50% and 95% prediction intervals. Vector control has been in place in Boffa West since 2016 (yellow bar), and the Ebola outbreak occurred between 2014 and 2016 (light purple background).

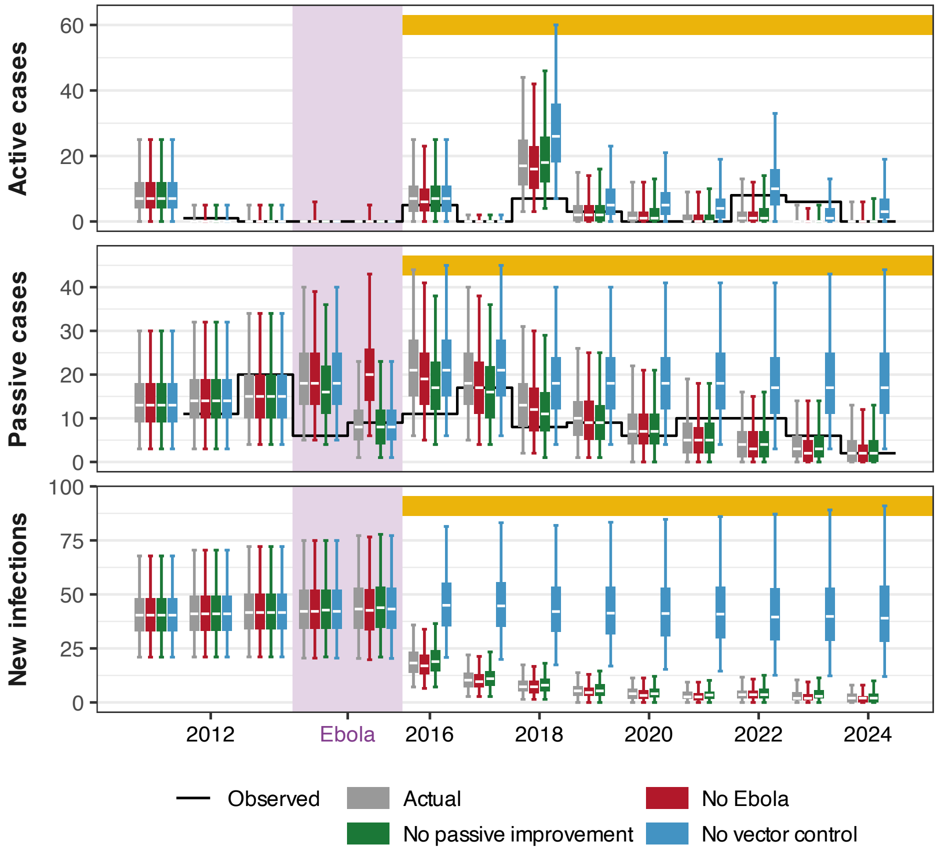

Fig F: Evolution of the number of new reported active and passive cases and new infections each year in Dubréka, according to the actual scenario (grey) and counterfactual scenarios: no Ebola outbreak (red), no improvement in passive screening (green), and no vector control (blue). Boxes and whiskers represent the median and the 50% and 95% prediction intervals. Vector control has been in place in Dubréka since 2016 (yellow bar), and the Ebola outbreak occurred between 2014 and 2016 (light purple background).

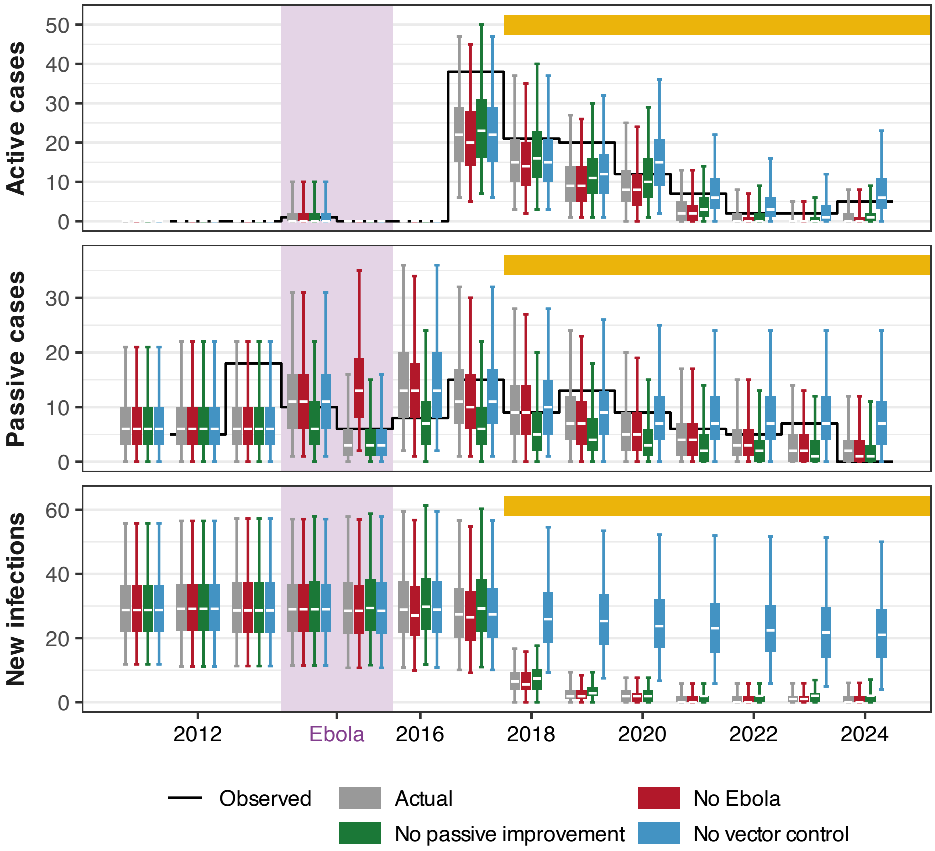

Fig G: Evolution of the number of new reported active and passive cases and new infections each year in Forécariah, according to the actual scenario (grey) and counterfactual scenarios: no Ebola outbreak (red), no improvement in passive screening (green), and no vector control (blue). Boxes and whiskers represent the median and the 50% and 95% prediction intervals. Vector control has been in place in Forécariah since 2018 (yellow bar), and the Ebola outbreak occurred between 2014 and 2016 (light purple background).

Table H: Summary of 2012–2024 transmission under the actual scenario and differences from this under three counterfactual scenarios.

| Focus | Variable | Scenario | | | | | | | | | | | |
|---|---|---|---|---|---|---|---|---|---|---|---|---|---|
| | | Actual | | | No Ebola | | | No PS improvement | | | No VC | | |
| | | | | | Diff. from Actual | | | Diff. from Actual | | | Diff. from Actual | | |
| | | med. | 95% CI | | med. | 95% CI | | med. | 95% CI | | med. | 95% CI | |
| Boffa West | New infections | 169 | 89 | 280 | -4 | -14 | 0 | 5 | 0 | 17 | 236 | 93 | 428 |
| | Cases | 183 | 106 | 264 | 7 | -1 | 17 | -12 | -29 | -2 | 122 | 54 | 207 |
| | Deaths | 87 | 43 | 158 | -11 | -23 | -4 | 14 | 3 | 31 | 35 | 12 | 76 |
| | DALYs | 3145 | 1581 | 5604 | -419 | -798 | -159 | 482 | 109 | 1087 | 1265 | 444 | 2708 |
| Boffa East | New infections | 62 | 24 | 125 | -2 | -8 | 0 | 3 | 0 | 13 | 356 | 135 | 761 |
| | Cases | 74 | 38 | 123 | 7 | 1 | 16 | -9 | -22 | -1 | 116 | 49 | 222 |
| | Deaths | 53 | 22 | 106 | -8 | -18 | -2 | 9 | 1 | 24 | 107 | 37 | 252 |
| | DALYs | 1813 | 762 | 3648 | -268 | -609 | -62 | 307 | 34 | 833 | 3710 | 1278 | 8671 |
| Dubréka | New infections | 226 | 121 | 386 | -3 | -12 | 0 | 5 | 0 | 18 | 317 | 140 | 624 |
| | Cases | 187 | 112 | 278 | 4 | -2 | 11 | -8 | -26 | 0 | 116 | 56 | 210 |
| | Deaths | 208 | 105 | 361 | -8 | -19 | -2 | 11 | 1 | 33 | 93 | 36 | 204 |
| | DALYs | 7409 | 3792 | 12784 | -300 | -658 | -73 | 390 | 39 | 1183 | 3334 | 1312 | 7214 |
| Forécariah | New infections | 188 | 92 | 354 | -4 | -13 | 0 | 7 | 0 | 21 | 148 | 55 | 305 |
| | Cases | 164 | 92 | 250 | 2 | -3 | 9 | -21 | -47 | -4 | 45 | 18 | 84 |
| | Deaths | 129 | 61 | 236 | -4 | -12 | 0 | 14 | 3 | 34 | 19 | 5 | 48 |
| | DALYs | 4883 | 2407 | 8735 | -160 | -450 | 0 | 540 | 118 | 1257 | 729 | 207 | 1772 |
| Country level totals: | | | | | | | | | | | | | |
| | New infections | 645 | 326 | 1145 | -13 | -47 | 0 | 20 | 0 | 69 | 1057 | 423 | 2118 |
| | Cases | 608 | 348 | 915 | 20 | -5 | 53 | -50 | -124 | -7 | 399 | 177 | 723 |
| | Deaths | 477 | 231 | 861 | -31 | -72 | -8 | 48 | 8 | 122 | 254 | 90 | 580 |
| | DALYs | 17250 | 8542 | 30771 | -1147 | -2515 | -294 | 1719 | 300 | 4360 | 9038 | 3241 | 20365 |

#### S1.6 Additional results

##### S1.6.1 Ensemble model joint posteriors of fitted parameters

Histograms of the 2 000 samples from the joint posterior distribution of the parameters of the ensemble model in each focus are presented in Figures H–K. If there was no variation in a parameter among the samples, that parameter was omitted from the plot – it is indicative that no samples were chosen from models in which that parameter was fitted.

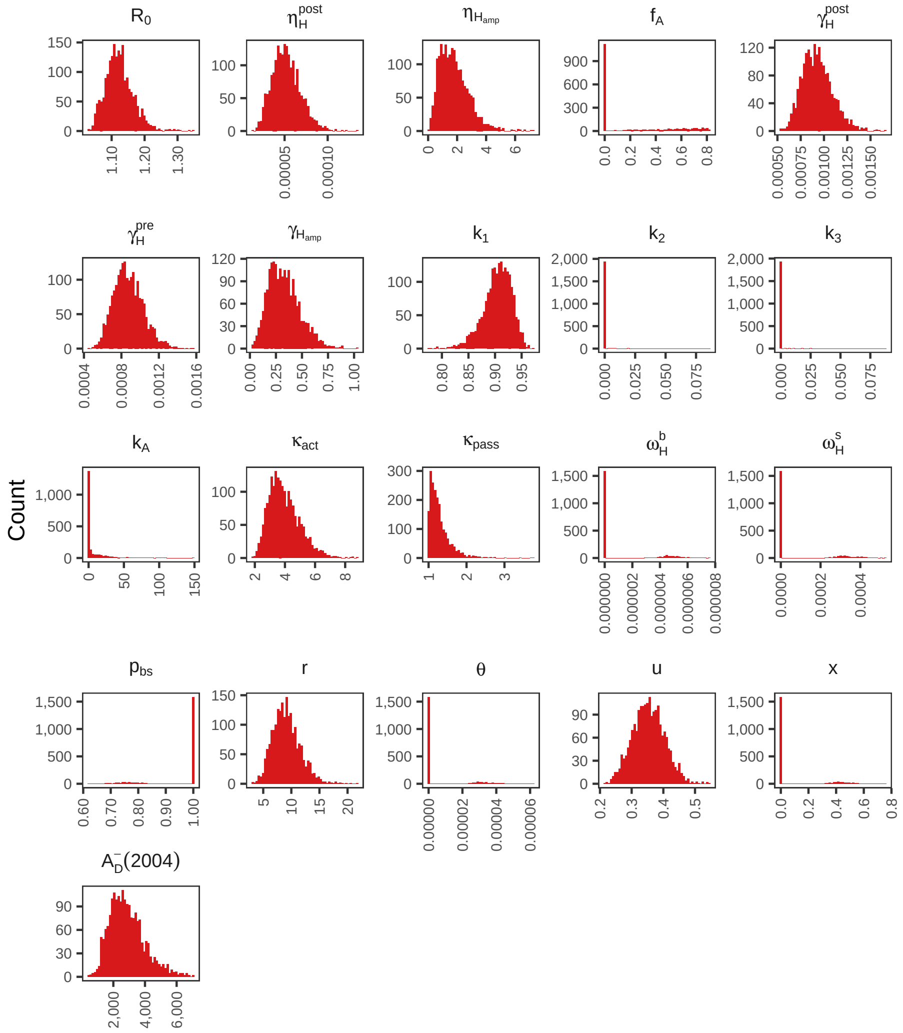

Fig H: Histograms of 2 000 samples from the joint posterior distribution of fitted parameters for the Boffa West ensemble model.

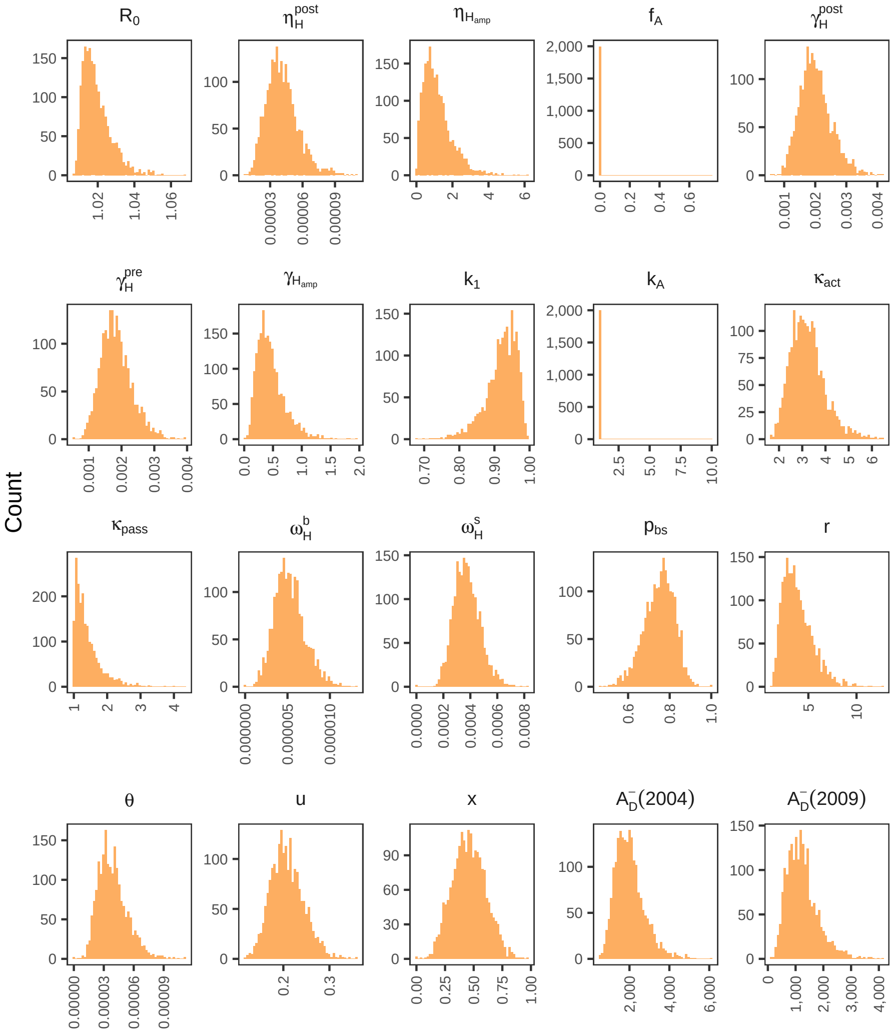

Fig I: Histograms of $2\,000$ samples from the joint posterior distribution of fitted parameters for the Boffa East ensemble model.

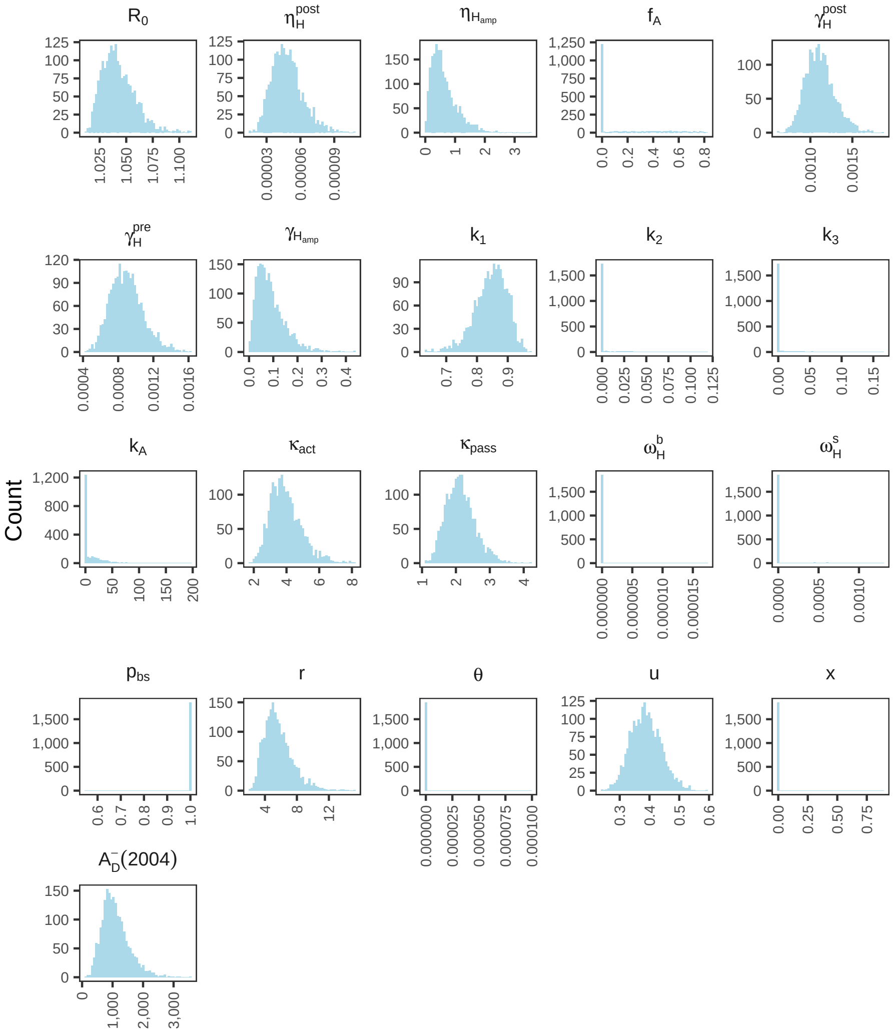

Fig J: Histograms of 2 000 samples from the joint posterior distribution of fitted parameters for the Dubréka ensemble model.

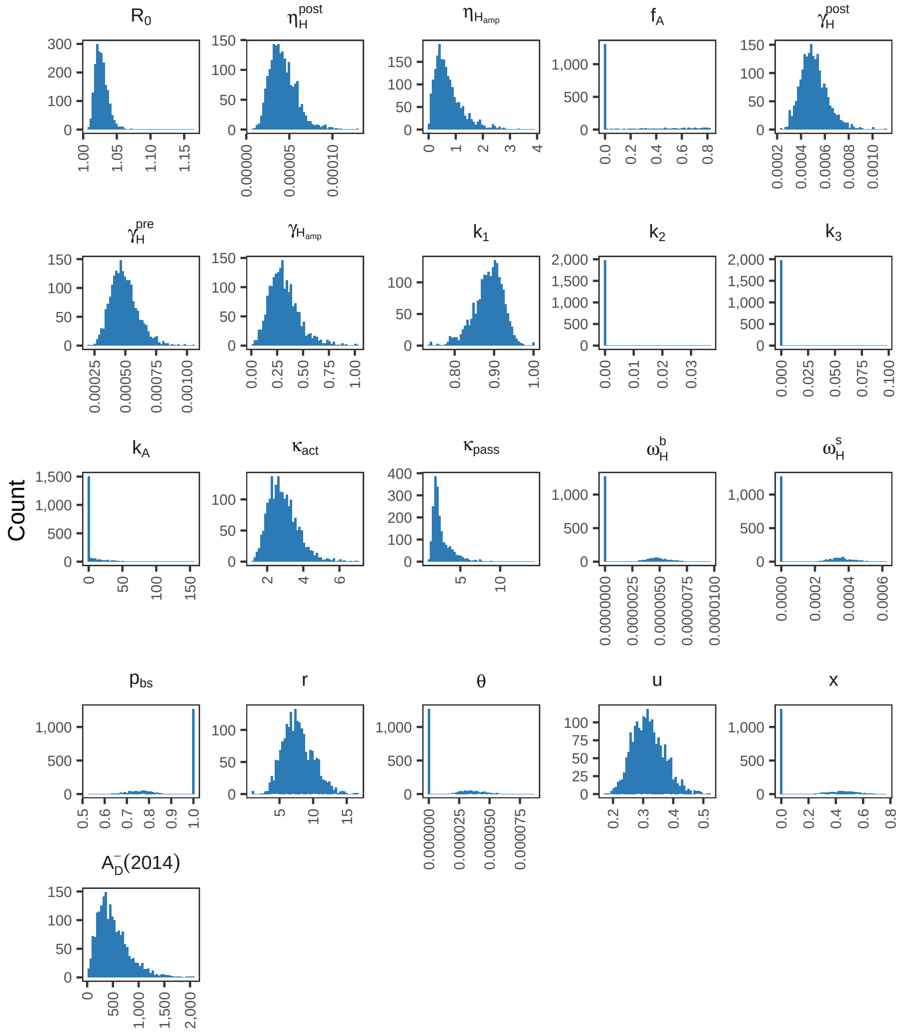

Fig K: Histograms of $2\,000$ samples from the joint posterior distribution of fitted parameters for the Forécariah ensemble model.

##### S1.6.2 Prevalence of infection

For the ensemble model, the prevalence of infection was calculated for $20\,000$ simulations across the fitted period, see Fig L. The within-year mean of the number of stage 1 and 2 individuals in all four human risk-by-AS participation groups ($\sum_{i=1}^{4} I_{1Hi}^{b} + I_{2Hi}^{b}$) plus the within-year mean of the number of individuals in the skin-only infection group ($I_H^s$) was used for the human prevalence of all infections, the within-year mean of skin-only infections ($I_H^s$) for the skin-only infection prevalence, and the within-year mean of $I_A$ for the animal prevalence. For vectors, we assume that infections may be detected anywhere within the tsetse (e.g. by molecular screening of the sampled tsetse which could include mid-gut infections), or specifically in the salivary glands (e.g. by dissection and microscopy) from where the infection can be passed on to host species. Therefore, both all infection and salivary gland infection prevalences were calculated, using the within-year mean of $E_{1V} + E_{2V} + E_{3V} + I_V$ or $I_V$.

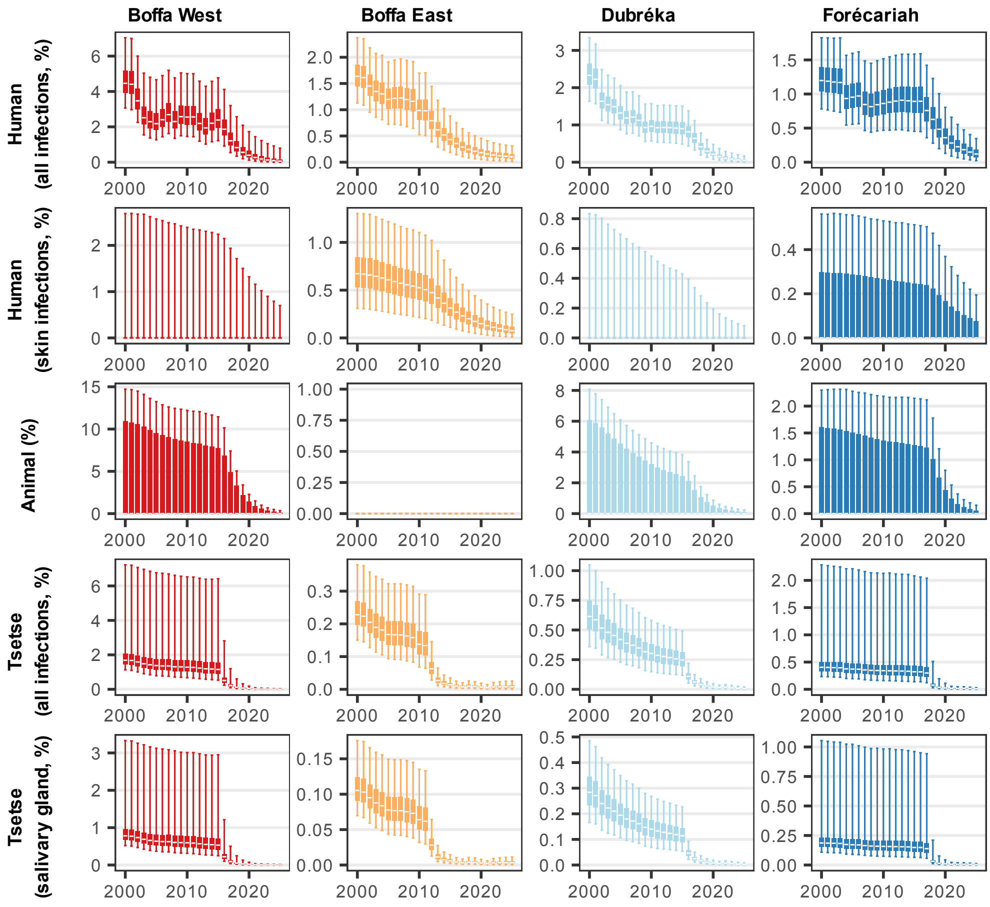

Fig L: Mean prevalence of infection within year as a percentage of the population. Human cases are considered across disease stages and population risk group. In vectors, the prevalence of all infections and infectious (salivary gland) infections is presented.

#### S1.7 PRIME-NTD criteria

Good modelling practices should include the Policy-Relevant Items for Reporting Models in Epidemiology of Neglected Tropical Diseases (PRIME-NTD) [59] table to address the five key principles of communication, quality and relevance of analyses. See Table I for how our epidemiological modelling part met the PRIME-NTD criteria in this paper.

Table I: **PRIME-NTD summary table.** We summarise how the NTD Modelling Consortium's "5 key principles of good modelling practice" have been met in the present study.

| What has been done to satisfy the principle? | Where in the manuscript is this described? |
|---|---|
| **1. Stakeholder engagement** | |
| This modelling study has been conducted alongside the national sleeping sickness control programme in Guinea and their collaborators. Numerous discussions were had to ensure the modellers produced meaningful outputs with policy relevance. This article is one piece of a larger, ongoing collaboration between the modelling group (HAT MEPP) and our partners. Regular meetings and occasional modelling workshops have been held over several years, where we discuss ongoing and planned strategies, historical data, and new data analyses. The original main text was written jointly in French before being translated into English. A draft version of this article was shared with the WHO HAT team, who review all of our modelling work which utilises the WHO HAT Atlas data. | Authorship list and acknowledgements |
| **2. Complete model documentation** | |
| Modelling code and documentation are available from the Open Science Framework (OSF) repository for this paper. The epidemiological model was fully described across the main text and this supplementary information. Previous versions of the model are described elsewhere [3, 5, 10, 11, 13, 14, 60–62]. | Methods in the main text, this SI, and on OSF |
| **3. Complete description of data used** | |
| The original human and tsetse data and how we aggregated the human data for fitting were described in the main text and this supplementary information. These data are included as supplementary information. | Methods section in the main text and this SI, also SI data files accompanying the paper and on OSF |
| **4. Communicating uncertainty** | |
| *Structural uncertainty*: We considered 9 model variants to incorporate the uncertainty of infection risk and participation in active screenings in humans as well as possible roles in transmission for animals and asymptomatic humans (skin-only infections). The model evidence, or marginal likelihood, for each model, was used to create our ensemble model for projections. | Methods sections in the main text and this SI. |
| *Parameter uncertainty*: We used samples from the joint posterior distributions of fitted parameters in the ensemble model generated from the model fitting process in our model simulations, plus stochastic variability also (10 realisations for each sample from the joint posterior of the ensemble model). | Model fitting procedure in this SI |
| *Prediction uncertainty*: We represent uncertainty in our results by providing box and whisker plots for simulated outcomes (median, 50% and 95% credible intervals), as well as histograms of the ensemble model posteriors in this SI. | Figure 3–5 in the main text, Figures D–L and Table H in this SI. |
| **5. Testable model outcomes** | |
| Our models were fitted to historical data. The results presented in this article alongside our open-source code and aggregated data files would allow replication and verification of our methodology and results. | Plots of fits to hisorical case data are presented in the main text. Code and appropriately formatted aggregated data are available from OSF. |

#### References

[1] Pandey A, Atkins KE, Bucheton B, Camara M, Aksoy S, Galvani AP, et al. Evaluating long-term effectiveness of sleeping sickness control measures in Guinea. Parasites & Vectors. 2015;8(1):550.

[2] Capewell P, Atkins K, Weir W, Jamonneau V, Camara M, Clucas C, et al. Resolving the apparent transmission paradox of African sleeping sickness. PLOS Biology. 2019 01;17:1-6.

[3] Antillon M, Huang CI, Sutherland SA, Crump RE, Brown PE, Bessell PR, et al. Cost-effectiveness of end-game strategies against sleeping sickness across the Democratic Republic of Congo. medRxiv. 2024. Available from: https://www.medrxiv.org/content/early/2024/03/30/2024.03.29.24305066.

[4] Kaba D, Koffi M, Kouakou L, N'Gouan EK, Djohan V, Courtin F, et al. Towards the sustainable elimination of human African trypanosomiasis in Côte d'Ivoire using an integrated approach. PLoS neglected tropical diseases. 2023;17(7):e0011514.

[5] Rock KS, Huang CI, Crump RE, Bessell PR, Brown PE, Tirados I, et al. Update of transmission modelling and projections of *gambiense* human African trypanosomiasis in the Mandoul focus, Chad. Infectious Diseases of Poverty. 2022;11(1):1-13.

[6] Camara M, Ouattara E, Duvignaud A, Migliani R, Camara O, Leno M, et al. Impact of the Ebola outbreak on Trypanosoma brucei gambiense infection medical activities in coastal Guinea, 2014-2015: A retrospective analysis from the Guinean national Human African Trypanosomiasis control program. PLOS Neglected Tropical Diseases. 2017 11;11(11):1-15. Available from: https://doi.org/10.1371/journal.pntd.0006060.

[7] Courtin F, Camara M, Rayaisse JB, Kagbadouno M, Dama E, Camara O, et al. Reducing Human-Tsetse Contact Significantly Enhances the Efficacy of Sleeping Sickness Active Screening Campaigns: A Promising Result in the Context of Elimination. PLoS Neglected Tropical Diseases. 2015 Sep;9(8):1-13.

[8] Courtin F, Jamonneau V, Camara M, Camara O, Coulibaly B, Diarra A, et al. A geographical approach to identify sleeping sickness risk factors in a mangrove ecosystem. Tropical Medicine & International Health. 2010;15(8):881-9. Available from: https://onlinelibrary.wiley.com/doi/abs/10.1111/j.1365-3156.2010.02559.x.

[9] Brinkhoff T. City Population. Guinea: Administrative Division; 2018. Accessed: 2021-02-03. https://www.citypopulation.de/en/guinea/admin/.

[10] Crump RE, Huang CI, Knock ES, Spencer SEF, Brown PE, Mwamba Miaka E, et al. Quantifying epidemiological drivers of *gambiense* human African Trypanosomiasis across the Democratic Republic of Congo. PLOS Computational Biology. 2021 01;17:1-23. Available from: https://doi.org/10.1371/journal.pcbi.1008532.

[11] Rock KS, Torr SJ, Lumbala C, Keeling MJ. Quantitative evaluation of the strategy to eliminate human African trypanosomiasis in the DRC. Parasites & Vectors. 2015;8(1):532.

[12] Rock KS, Torr SJ, Lumbala C, Keeling MJ. Predicting the impact of intervention strategies for sleeping sickness in two high-endemicity health zones of the Democratic Republic of Congo. PLoS Neglected Tropical Diseases. 2017;11:e0005162.

[13] Mahamat MH, Peka M, Rayaisse Jb, Rock KS, Toko MA, Darnas J, et al. Adding tsetse control to medical activities contributes to decreasing transmission of sleeping sickness in the Mandoul focus (Chad). PLoS Neglected Tropical Diseases. 2017;11(7):e0005792.

[14] Crump RE, Aliee M, Sutherland SA, Huang CI, Crowley EH, Spencer SEF, et al. Modelling timelines to elimination of sleeping sickness in the Democratic Republic of Congo, accounting for possible cryptic human and animal transmission. Parasites & Vectors. 2024 Aug;17(1):332. Available from: https://doi.org/10.1186/s13071-024-06404-4.

[15] Davis CN, Crump RE, Sutherland SA, Spencer SE, Corbella A, Chansy S, et al. Comparison of stochastic and deterministic models for gambiense sleeping sickness at different spatial scales: A health area analysis in the DRC. PLOS Computational Biology. 2024;20(4):e1011993.

[16] The World Bank. Life expectancy at birth, total (years) - Guinea; 2024. Accessed: 24 July 2024. Available from: https://data.worldbank.org/indicator/SP.DYN.LE00.IN?locations=GN.

[17] Rogers DJ. A general model for the African trypanosomiases. Parasitology. 1988;97:193-212.

[18] Checchi F, Filipe JAN, Haydon DT, Chandramohan D, Chappuis F. Estimates of the duration of the early and late stage of gambiense sleeping sickness. BMC Infectious Diseases. 2008;8(1):16-6.

[19] Checchi F, Funk S, Chandramohan D, Haydon DT, Chappuis F. Updated estimate of the duration of the meningo-encephalitic stage in *gambiense* human African trypanosomiasis. BMC Res Notes. 2015;8(1):292.

[20] World Health Organisation. Guidelines for the treatment of human African trypanosomiasis; 2024. Accessed: 7 August 2024. Available from: https://www.who.int/publications/i/item/9789240096035#:~:text=The%20WHO%20interim%20guidelines%20for,for%20treatment%20of%20rhodesiense%20HAT.

[21] Gouteux JP, Jarry M. Tsetse flies, biodiversity and the control of sleeping sickness. Structure of a Glossina guild in southwest Côte d'Ivoire. Acta Oecologica. 1998;19(5):453-71.

[22] Hargrove J. Extinction probabilities and times to extinction for populations of tsetse flies Glossina spp.(Diptera: Glossinidae) subjected to various control measures. Bulletin of Entomological Research. 2005;95(1):13-21.

[23] Davis S, Aksoy S, Galvani AP. A global sensitivity analysis for African sleeping sickness. Parasitology. 2010;138(04):516-26.

[24] Ravel S, Grebaut P, Cuisance D, Cuny G. Monitoring the developmental status of *Trypanosoma brucei gambiense* in the tsetse fly by means of PCR analysis of anal and saliva drops. Acta Tropica. 2003;88(2):161-5.

[25] World Health Organisation. Control and surveillance of human African trypanosomiasis. World Health Organisation; 2013. 984.

[26] Kubi C, van den Abbeele J, de Deken R, T MARCOTTY T, Dorny P, P van den Bossche P. The effect of starvation on the susceptibility of teneral and non-teneral tsetse flies to trypanosome infection. Medical and Veterinary Entomology. 2021;20:388-92.

[27] Clausen PH, Adeyemi I, Bauer B, Breloeer M, Salchow F, Staak C. Host preferences of tsetse (Diptera: Glossinidae) based on bloodmeal identifications. Medical and Veterinary Entomology. 1998;12(2):169-80.

[28] Pépin J, Guern C, Mercier D, Moore P. Utilisation du Testryp CATT pour le dépistage de la trypanosomiase à Nioki, Zaïre [Use of the CATT Testryp in screening for trypanosomiasis in Nioki, Zaire]. Annales de la Societe belge de medecine tropicale. 1986;66(3):213-24.

[29] Noireau F, Lemesre JL, Nzoukoudi MY, Louembet MT, Gouteux JP, Frezil JL. Serodiagnosis of sleeping sickness in the Republic of the Congo: Comparison of indirect immunofluorescent antibody test and card agglutination test. Transactions of The Royal Society of Tropical Medicine and Hygiene. 1988 03;82(2):237-40. Available from: https://doi.org/10.1016/0035-9203(88)90430-0.

[30] Enyaru JCK, Matovu E, Akol M, Sebikali C, Kyambadde J, Schmidt C, et al. Parasitological detection of Trypanosoma brucei gambiense in serologically negative sleeping-sickness suspects from north-western Uganda. Annals of Tropical Medicine & Parasitology. 1998;92(8):845-50. PMID: 10396344. Available from: https://doi.org/10.1080/00034983.1998.11813349.

[31] Jamonneau V, Truc P, Garcia A, Magnus E, Büscher P. Preliminary evaluation of LATEX/T. b. gambiense and alternative versions of CATT/T. b. gambiense for the serodiagnosis of Human African Trypanosomiasis of a population at risk in Côte d'Ivoire: considerations for mass-screening. Acta Tropica. 2000;76(2):175-83. Available from: https://www.sciencedirect.com/science/article/pii/S0001706X00000954.

[32] Truc P, Lejon V, Magnus E, Jamonneau V, Nangouma A, Verloo D, et al. Evaluation of the micro-CATT, CATT/Trypanosoma brucei gambiense, and LATEX/T b gambiense methods for serodiagnosis and surveillance of human African trypanosomiasis in West and Central Africa. Bulletin of the World Health Organisation. 2002;80(11):882-6. PMID: 12481210.

[33] Magnus E, Lejon V, Bayon D, Buyse D, Simarro P, Verloo D, et al. Evaluation of an EDTA version of CATT/Trypanosoma bruceigambiense for serological screening of human blood samples. Acta Tropica. 2002;81(1):7-12. Available from: https://www.sciencedirect.com/science/article/pii/S0001706X0100184X.

[34] Penchenier L, Grébaut P, Njokou F, Eyenga VE, Büscher P. Evaluation of LATEX/T.b.gambiense for mass screening of Trypanosoma brucei gambiense sleeping sickness in Central Africa. Acta Tropica. 2003;85(1):31-7. Available from: https://www.sciencedirect.com/science/article/pii/S0001706X02002322.

[35] Inojosa WO, Augusto I, Bisoffi Z, Josenado T, Abel PM, Stich A, et al. Diagnosing human African trypanosomiasis in Angola using a card agglutination test: observational study of active and passive case finding strategies. BMJ. 2006;332(7556):1479. Available from: https://www.bmj.com/content/332/7556/1479.

[36] Büscher P, Mertens P, Leclipteux T, Gilleman Q, Jacquet D, Mumba-Ngoyi D, et al. Sensitivity and specificity of HAT Sero-¡em¿K¡/em¿-SeT, a rapid diagnostic test for serodiagnosis of sleeping sickness caused by ¡em¿Trypanosoma brucei gambiense¡/em¿: a case-control study. The Lancet Global Health. 2014 Jun;2(6):e359-63. Available from: https://doi.org/10.1016/S2214-109X(14)70203-7.

[37] Bisser S, Lumbala C, Nguertoum E, Kande V, Flevaud L, Vatunga G, et al. Sensitivity and Specificity of a Prototype Rapid Diagnostic Test for the Detection of Trypanosoma brucei gambiense Infection: A Multi-centric Prospective Study. PLOS Neglected Tropical Diseases. 2016 04;10(4):1-16. Available from: https://doi.org/10.1371/journal.pntd.0004608.

[38] Lumbala C, Bessell PR, Lutumba P, Baloji S, Biéler S, Ndung'u JM. Performance of the SD BIOLINE® HAT rapid test in various diagnostic algorithms for gambiense human African trypanosomiasis in the Democratic Republic of the Congo. PLOS ONE. 2017 07;12(7):1-17. Available from: https://doi.org/10.1371/journal.pone.0180555.

[39] Lumbala C, Biéler S, Kayembe S, Makabuza J, Ongarello S, Ndung'u JM. Prospective evaluation of a rapid diagnostic test for Trypanosoma brucei gambiense infection developed using recombinant antigens. PLOS Neglected Tropical Diseases. 2018 03;12(3):1-20. Available from: https://doi.org/10.1371/journal.pntd.0006386.

[40] N'Djetchi MK, Camara O, Koffi M, Camara M, Kaba D, Kaboré J, et al. Specificity of serological screening tests and reference laboratory tests to diagnose gambiense human African trypanosomiasis: a prospective clinical performance study. Infectious Diseases of Poverty. 2024 Jul;13(1):53. Available from: https://doi.org/10.1186/s40249-024-01220-5.

[41] Tablado Alonso S, Biéler S, Inocêncio Da Luz R, Verlé P, Büscher P, Hasker E. Phase I evaluation of the Abbott Bioline HAT 2.0, a rapid diagnostic test for Human African Trypanosomiasis based on recombinant antigens. In: 36TH General Conference of the International Scientific Council for Trypanosomiasis Research and Control; 2023. .

[42] Tablado Alonso S, Inocêncio Da Luz R, Verlé P, Büscher P, Mumba Ngoyi D, Mwamba Miaka D, et al. Assessment of recombinant HAT-RDT specificity. In: 36TH General Conference of the International Scientific Council for Trypanosomiasis Research and Control; 2023. .

[43] Lumsden WHR, Kimber CD, Dukes P, Haller L, Stanghellini A, Duvallet G. Field diagnosis of sleeping sickness in the Ivory Coast. I. Comparison of the miniature anion-exchange/centrifugation technique with other protozoological methods. Transactions of the Royal Society of Tropical Medicine and Hygiene. 1981;75(2):242-50. Available from: https://www.sciencedirect.com/science/article/pii/0035920381903266.

[44] Dukes P, Rickman L, Killick-Kendrick R, Kakoma I, Wurapa F, de Raadt P, et al. A field comparison of seven diagnostic techniques for human trypanosomiasis in the Luangwa Valley, Zambia. Tropenmedizin und Parasitologie. 1984 Sep;35(3):141-7. PMID: 6208660.

[45] Miézan T, MEDA A, DOUA F, CATTAND P. Evaluation des techniques parasitologiques utilisées dans le diagnostic de la trypanosome humaine à Trypanosoma gambiense en Côte-d'Ivoire. Bulletin de la Société de pathologie exotique. 1994;87(2):101-4.

[46] Truc P, Bailey JW, Doua F, Laveissière C, Godfrey DG. A comparison of parasitological methods for the diagnosis of gambian trypanosomiasis in an area of low endemicity in Côte d'Ivoire. Transactions of the Royal Society of Tropical Medicine and Hygiene. 1994;88(4):419-21. Available from: https://www.sciencedirect.com/science/article/pii/0035920394904103.

[47] Lutumba P, Robays J, Miaka C, Kande V, Mumba D, Büscher P, et al. [Validity, cost and feasibility of the mAECT and CTC confirmation tests after diagnosis of African of sleeping sickness]. Tropical medicine

& international health. 2006 April;11(4):470-8. Available from: https://onlinelibrary.wiley.com/doi/pdfdirect/10.1111/j.1365-3156.2006.01591.x.

[48] Haines LR. Examining the tsetse teneral phenomenon and permissiveness to trypanosome infection. Frontiers in cellular and infection microbiology. 2013;3:84.

[49] Huang CI, Crump RE, Brown PE, Spencer SE, Miaka EM, Shampa C, et al. Identifying regions for enhanced control of *gambiense* sleeping sickness in the Democratic Republic of Congo. Nature communications. 2022;13(1):1-11.

[50] Tirados I, Esterhuizen J, Kovacic V, Mangwiro TNC, Vale GA, Hastings I, et al. Tsetse Control and Gambian Sleeping Sickness; Implications for Control Strategy. PLOS Neglected Tropical Diseases. 2015;9:1-22.

[51] Tirados I, Hope A, Selby R, Mpembele F, Miaka EM, Boelaert M, et al. Impact of tiny targets on *Glossina fuscipes quanzensis*, the primary vector of human African trypanosomiasis in the Democratic Republic of the Congo. PLOS Neglected Tropical Diseases. 2020;14:1-20.

[52] Kaba D, Djohan V, Berté D, Ta BTD, Selby R, Kouadio KADM, et al. Use of vector control to protect people from sleeping sickness in the focus of Bonon (Côte d'Ivoire). PLoS Neglected Tropical Diseases. 2021;15(6):e0009404.

[53] Storn R, Price K. Differential Evolution – A Simple and Efficient Heuristic for global Optimization over Continuous Spaces. Journal of Global Optimization. 1997 Dec;11(4):341-59. Available from: https://doi.org/10.1023/A:1008202821328.

[54] Conceicao ELT. DEoptimR: Differential Evolution Optimization in Pure R; 2025. R package version 1.1-4. Available from: https://CRAN.R-project.org/package=DEoptimR.

[55] Brest J, Greiner S, Boskovic B, Mernik M, Zumer V. Self-Adapting Control Parameters in Differential Evolution: A Comparative Study on Numerical Benchmark Problems. IEEE Transactions on Evolutionary Computation. 2006;10(6):646-57.

[56] Touloupou P, Alzahrani N, Neal P, Spencer SEF, McKinley TJ. Efficient Model Comparison Techniques for Models Requiring Large Scale Data Augmentation. Bayesian Anal. 2018 06;13(2):437-59. Available from: https://doi.org/10.1214/17-BA1057.

[57] Hesterberg T. Weighted Average Importance Sampling and Defensive Mixture Distributions. Technometrics. 1995;37(2):185-94. Available from: https://www.jstor.org/stable/1269620.

[58] Sunnucks R, Davis EL, Rock KS. Methods for Reproducible Comparison of Strategies in Stochastic Modelling. medRxiv. 2025. Available from: https://www.medrxiv.org/content/early/2025/10/10/2025.10.09.25337145.

[59] Behrend MR, Basáñez MG, Hamley JI, Porco TC, Stolk WA, Walker M, et al. Modelling for policy: the five principles of the Neglected Tropical Diseases Modelling Consortium. PLoS neglected tropical diseases. 2020;14(4):e0008033.

[60] Rock KS, Pandey A, Ndeffo-Mbah ML, Atkins KE, Lumbala C, Galvani A, et al. Data-driven models to predict the elimination of sleeping sickness in former Equateur province of DRC. Epidemics. 2017;18:101-12.

[61] Crump RE, Huang CI, Spencer SEF, Brown PE, Shampa C, Miaka EM, et al. Modelling to infer the role of animals in gambiense human African trypanosomiasis transmission and elimination in the DRC. PLoS Neglected Tropical Diseases. 2022;16(7):1-23.

[62] Huang CI, Crump RE, Brown PE, Spencer SEF, Miaka EM, Shampa C, et al. Identifying regions for enhanced control of gambiense sleeping sickness in the Democratic Republic of Congo. Nature Communications. 2022 Mar;13(1):1448. Available from: https://doi.org/10.1038/s41467-022-29192-w.
